# Ophthalmic AI With Just Ten Examples: Comparing Multimodal Large Language Models, Vision Foundation Models, and Vision-Language Models

**DOI:** 10.64898/2026.10.30.26366929

**Authors:** Pratyush Ramakrishna, Jack Strand, Scott Song, Mahsa B. Tavakoli, Samuel Zetumer, Shahriyar Majidi, Yu Jiang, Mengxi Shen, Alessandro Berni, Giovanni Gregori, Philip J. Rosenfeld, Michael Kass, Mae Gordon, Cecilia S. Lee, Yuka Kihara, Aaron Y. Lee

## Abstract

**Purpose:** To compare multimodal large language models (MLLMs), vision foundation models (VFMs), and contrastive vision-language models (VLMs) for ophthalmic image interpretation with no labeled examples (zero-shot) or as few as 10 labeled examples per class (10-shot).

**Design:** Retrospective benchmarking study.

**Subjects:** 4 public datasets (UCSD, IDRiD, REFUGE, ASOCT-MSDC) and 3 private datasets (OHTS, BPEI-DLS, WashU-OCT) of anterior-and posterior-segment ophthalmic imaging.

**Methods:** Nine AI models were evaluated across 3 architectural families: MLLMs (Qwen3.8-27B, GLM-5.3-Flash, Gemma-4-31B-it), VFMs (DINOv3, RETFound, VisionFM), and VLMs (CLIP, EyeCLIP, SigLIP2). Models were assessed on disease classification, disease risk and severity grading, anatomical assessment, and lesion localization in fundus and OCT imaging. MLLMs were also compared with a 3D VFM (OCTCube-M) for whole-volume OCT classification. MLLMs were evaluated zero-shot and 10-shot, without additional training. Vision models (VFMs and VLMs) were adapted by linear probing and fine-tuning on the same 10-shot set and on the full training dataset (FD).

**Main Outcome Measures:** Disease classification performance (macro-averaged F1, AUROC, quadratic weighted kappa [QWK]), anatomical localization accuracy, and cross-dataset generalization.

**Results:** For retinal OCT disease classification, 10-shot MLLMs achieved a macro-F1 of 0.94, versus 0.59 (10-shot) and 0.96 (FD) for vision models. For diabetic retinopathy and macular edema grading, 10-shot MLLMs achieved QWKs of 0.69 and 0.73, versus 0.48 and 0.57 (10-shot) and 0.72 and 0.79 (FD) for vision models. For lesion localization, 10-shot MLLMs achieved an F1 of 0.75, versus 0.70 (10-shot) and 0.86 (FD) for vision models. For vertical cup-to-disc ratio estimation (OHTS), zero-shot MLLMs achieved a mean absolute error of 0.14, versus 0.21 (10-shot) for vision models. For whole-volume OCT classification (WashU-OCT), zero-shot MLLMs achieved a macro-F1 of 0.71, versus 0.50 (10-shot) and 0.82 (FD) for vision models.

**Conclusions:** Open-weight MLLMs given 10 labeled examples per class outperformed VFMs and VLMs trained on the same examples in most tasks and approached the performance of fully trained VFMs and VLMs in selected disease classification and grading tasks. MLLMs also demonstrated promising zero-shot performance on non-public clinical datasets without task-specific training. By reducing the need for large labeled datasets and task-specific model adaptation, MLLMs could broaden who can develop and validate ophthalmic AI.

## INTRODUCTION

Deep learning has enabled accurate detection of several eye diseases from color fundus photographs (CFPs) and optical coherence tomography (OCT), including diabetic retinopathy (DR), age-related macular degeneration (AMD) and glaucomatous optic neuropathy.^1–6^ However, developing and adapting these systems have traditionally required large amounts of labeled data.^6,7^ Several prominent systems, for example, were developed using tens of thousands to more than 100,000 expert-graded fundus photographs^1,2^ or thousands of labeled OCT volumes.^6^ Expert annotation is time-consuming,^7^ grader disagreement makes reference standards imperfect,^8^ and public ophthalmic datasets are geographically concentrated, with incomplete reporting of patient demographics and labeling methods.^9^ In addition, performance can vary substantially across algorithms and screening sites, which supports local validation before deployment.^10^

Vision foundation models (VFMs), such as RETFound and VisionFM, may reduce the need for new task-specific annotations by learning from large image collections before being adapted to specific clinical tasks.^11–14^ RETFound, for example, was pretrained on 1.6 million unlabeled retinal images and, when adapted to ophthalmic tasks, outperformed comparison models while needing fewer labeled images.^13^ Contrastive vision-language models (VLMs), such as CLIP, instead learn to match images with paired text, which allows them to classify an image by comparing it with written descriptions of each disease category.^15,16^ In ophthalmology, VLMs such as FLAIR and EyeCLIP have performed well when adapted with few labeled images, following pretraining on large ophthalmic imaging datasets.^17,18^

Generative multimodal large language models (MLLMs), a class of models distinct from VFMs and VLMs,^19^ may reduce labeled data requirements even further. By connecting a vision encoder to a large language model, an MLLM can follow written instructions about an image and respond in free text.^20,21^ MLLMs can learn a new task from a few labeled examples in the prompt without retraining, an approach known as in-context learning.^22,23^ General-purpose MLLMs such as GPT-4o and Gemini can interpret ophthalmic images, although performance has varied by disease, imaging type and task,^24–27^ and adding examples to the prompt has improved it in some evaluations but not in others.^28,29^ Additionally, many of these evaluations used proprietary models accessed through the developer’s cloud service.^23,24,27,28^ Sending images containing protected health information to an external cloud service for processing on behalf of a covered institution requires a business associate agreement and other applicable safeguards under the Health Insurance Portability and Accountability Act (HIPAA).^30,31^ Developers may also change or discontinue access to proprietary models after validation.^32^ Open-weight MLLMs, whose model weights are publicly released, can instead run entirely on an institution’s own hardware.^21,32^ Such locally deployed models keep both patient data and the model under institutional control, which may ease privacy and regulatory concerns about their clinical use.^21,32^

Evidence remains limited on whether locally deployed MLLMs match VFMs and VLMs across diverse ophthalmic tasks and datasets. Recent benchmarking studies have reported mixed findings on the value of ophthalmology-specific pretraining, with general-purpose VFMs matching or outperforming retina-specific VFMs on some tasks,^33,34^ and retina-specific VFMs retaining an advantage on others.^35^ FOCUS (Foundation Ophthalmic Cross-Dataset Understanding under Shift) further demonstrated that no model family consistently performed best across tasks and datasets, but evaluated only color fundus photograph (CFP) classification.^19^ Prior exposure of proprietary models to public imaging datasets can also be difficult to exclude. We therefore sought to benchmark local, open-weight MLLMs against VLMs and VFMs across a wide variety of ophthalmic image analysis tasks on both public and private datasets.

## METHODS

This study was conducted in accordance with the tenets of the Declaration of Helsinki and was approved by the Institutional Review Board (IRB) of Washington University in St. Louis. Due to the retrospective nature of the study, which involved the analysis of previously collected clinical data, the IRB waived the requirement for informed consent. All analyses were performed on a secured HIPAA-compliant local GPU server.

We conducted a benchmark study to evaluate the performance of emerging generative MLLMs relative to established vision models (VFMs and VLMs) for ophthalmic image interpretation tasks across 7 imaging datasets (4 public and 3 private). Tasks included disease classification, disease risk and severity grading, anatomical measurements, and lesion localization in CFPs, posterior-segment OCT, and anterior-segment OCT (AS-OCT), as well as multi-image classification of real-world clinical OCT volumes.

A primary aim of this study was to assess how model performance changed with the number of labeled examples, such as images with known diagnoses, from which a model can learn a new task. First, models were evaluated without any labeled examples from the task’s dataset (zero-shot evaluation). Second, models were evaluated after being given 10 labeled examples per class (10-shot evaluation). To ensure a fair comparison, the examples provided to MLLMs were the same examples provided to vision models, for each task and random seed (42–46). For a complete evaluation, vision models were also evaluated after being given all available labeled examples (full-data adaptation). This was not performed for MLLMs, as current models cannot accept thousands of example images within a single prompt.

MLLMs and vision models learned new tasks from these labeled examples in different ways. MLLMs received written task instructions and the labeled examples within a prompt, from which they generated predictions without any additional training, an approach known as in-context learning. In contrast, vision models underwent additional training on the labeled examples in one of three ways, which differ in how much of the pretrained model is changed: linear probing, in which the model’s internal weights are kept fixed and only a simple final classification layer is trained; partial fine-tuning, in which the weights of the last few layers are also adjusted; and full fine-tuning, in which all weights are adjusted.

### Ophthalmic Image Datasets

Seven datasets spanning three modalities were used (Supplementary Table S1): 1) University of California San Diego (UCSD) OCT dataset for four-class disease classification,^5^ the 2) Indian Diabetic Retinopathy Image Dataset (IDRiD) for diabetic retinopathy severity (grades 0–4) and diabetic macular edema risk (grades 0–2) grading,^36,37^ the 3) Retinal Fundus Glaucoma Challenge (REFUGE) dataset for glaucoma classification with optic disc and cup localization, alongside the 4) Ocular Hypertension Treatment Study (OHTS) dataset for external validation,^38,39^ the 5) AS-OCT Multi-Structure and Disease Classification Dataset (ASOCT-MSDC) for angle classification and scleral-spur localization,^40^ an 6) institutional spectral-domain retinal OCT dataset (BPEI-DLS) with expert-annotated drusen and double-layer signs (DLS) for band-level lesion localization, and a 7) private, institutionally-curated EHR-linked clinical OCT volumes (WashU-OCT) for whole-volume OCT classification.

### Evaluated Model Families

We evaluated nine AI models across three distinct architectural families. 1) VFMs included the general-purpose DINOv3,^41^ and ophthalmic-specific RETFound^13^ and VisionFM,^14^ which learn general visual features from large image collections and are adapted to specific tasks; 2) Contrastive VLMs included general purpose CLIP^15^ and SigLIP2^42^ as well as ophthalmic-specific EyeCLIP,^18^ which learn to align images with text descriptions; 3) Generative MLLMs included the general purpose Qwen3.8-27B (27 billion parameters),^43^ GLM-5.3-Flash (320 billion total parameters, with 18 billion activated per token; mixture-of-experts architecture),^44^ and Gemma-4-31B-it (31 billion parameters),^45^ which can process multiple modalities including images alongside textual prompts to generate natural-language responses.

Model checkpoints, input resolutions, and inference configurations are provided in Supplementary Table S2-S3. Additional training and prompting details are provided in Appendix 2-3. Few-shot supervised adaptation used 10 training epochs, except for localization tasks, which used 30 epochs (Supplementary Table S3).

### Dataset-Specific Protocols

The tasks and primary evaluation metrics for each dataset are summarized below; WashU-OCT is described separately in the next section.

#### UCSD

We evaluated four-class B-scan classification (normal, choroidal neovascularization [CNV], diabetic macular edema [DME], and drusen), with macro-averaged F1 as the primary metric.

#### IDRiD

We evaluated five-grade DR severity and three-grade DME risk classification, with quadratic-weighted kappa as the primary metric.

#### REFUGE

We evaluated vertical cup-to-disc ratio (vCDR) estimation from predicted optic disc and cup boundaries, and glaucoma discrimination using vCDR as a continuous score, with mean absolute error (MAE) and the area under the receiver operating characteristic curve (AUROC) as the respective primary metrics.

#### OHTS

We evaluated cross-dataset generalization of vCDR estimation, with MAE as the primary metric; few-shot demonstrations were derived exclusively from REFUGE. To assess demographic robustness, vCDR error on OHTS was also analyzed across age, sex, and race subgroups.

#### ASOCT-MSDC

We evaluated open-versus closed-angle classification and scleral-spur landmark localization, with macro-averaged F1 and Euclidean distance error as the respective primary metrics.

#### BPEI-DLS

We evaluated drusen and DLS localization, with band-level F1 as the primary metric. Each B-scan was divided into eight vertical bands, and bands containing at least four annotated lesion pixels were considered positive. Vision models predicted band labels through supervised training, whereas MLLMs identified lesion-containing bands among numbered regions using Set-of-Marks-inspired prompting.^46^

To assess whether predictions relied on clinically relevant image regions, we performed occlusion sensitivity analyses for the UCSD and IDRiD datasets. Each input image was divided into an 8 × 8 grid, and each of the 64 cells was masked in turn. The change in the model’s score for the target class was recorded for each masked grid, producing a map of the image regions driving each prediction.

### Volumetric Extension: EHR-Linked Clinical OCT

From the institutional Spectralis archive at Washington University in St Louis (199,486 imaging series from 19,588 patients; 2013–2026), we constructed the Washington University Clinical OCT Cohort (WashU-OCT), an enriched, single-disease cohort of 2,031 patients, one OCT volume each, with EHR-derived reference labels: 752 AMD, 627 DME, 353 epiretinal membrane (ERM), and 299 healthy. Labels were assigned from eye-specific historical ICD-10 diagnoses and refined by rule-based screening of clinical notes. The dataset was partitioned at the patient level into a training set (n = 426), a validation set (n = 183), and a held-out test set (n = 1,422). Five 10-shot sets were constructed by randomly sampling 10 patients per class from the training set, using a different random seed for each set (seeds 42–46). Full details of cohort construction are provided in Appendix 3 (WashU OCT Cohort Construction).

Three MLLMs were evaluated in the zero-shot setting, with each OCT volume represented by 25 B-scans provided together in a single inference request. OCTCube-M, a pretrained three-dimensional OCT foundation model, served as the supervised comparator.^47^ OCTCube-M was adapted by linear probing and partial fine-tuning using either a 10-shot set (10 labeled volumes per class) or the full training set, with checkpoints selected on the validation set. All models were evaluated on the same held-out test set.

### Task-Specific Statistics

For disease classification tasks, macro-averaged F1 was the primary metric, supplemented by balanced accuracy and per-class metrics; for glaucoma discrimination, AUROC was the primary metric. For grading classification tasks, quadratic-weighted kappa was the primary metric, supplemented by adjacent-grade accuracy. For lesion localization, band-level F1 was the primary metric, supplemented by balanced accuracy. For landmark localization, Euclidean distance error was the primary metric. For anatomical measurement, vCDR mean absolute error (MAE) was the primary metric. For few-shot evaluation, results were reported as the mean ± SD across random seeds, with bootstrap 95% CIs for key comparisons. For MLLMs, unparseable responses were logged and reported separately as an invalid-response rate.

## RESULTS

We evaluated ophthalmic image interpretation across three model families: VFMs, contrastive VLMs, and generative MLLMs. Tasks included disease classification, disease risk and severity grading, anatomical measurements, anatomical landmark and lesion localization, cross-dataset generalization, and whole-volume OCT classification, across multiple imaging modalities.

Models were compared with no labeled examples (zero-shot), with the same 10 labeled examples per class (10-shot), and, for vision models, with all available labeled data (full-data adaptation). Occlusion sensitivity analyses assessed which image regions influenced predictions. Figure 1 illustrates the study design, Figure 2 summarizes performance comparisons, and Figure 3 presents localization and interpretability analyses, and Figure 4 compares performance across labeled example budgets (A, B), cross-dataset generalization (C), and the volumetric evaluation (D). Appendix 1 provide detailed results across models and supervision settings, including 5-shot experiments and partial fine-tuning, with 95% confidence intervals.

**Figure 1.**
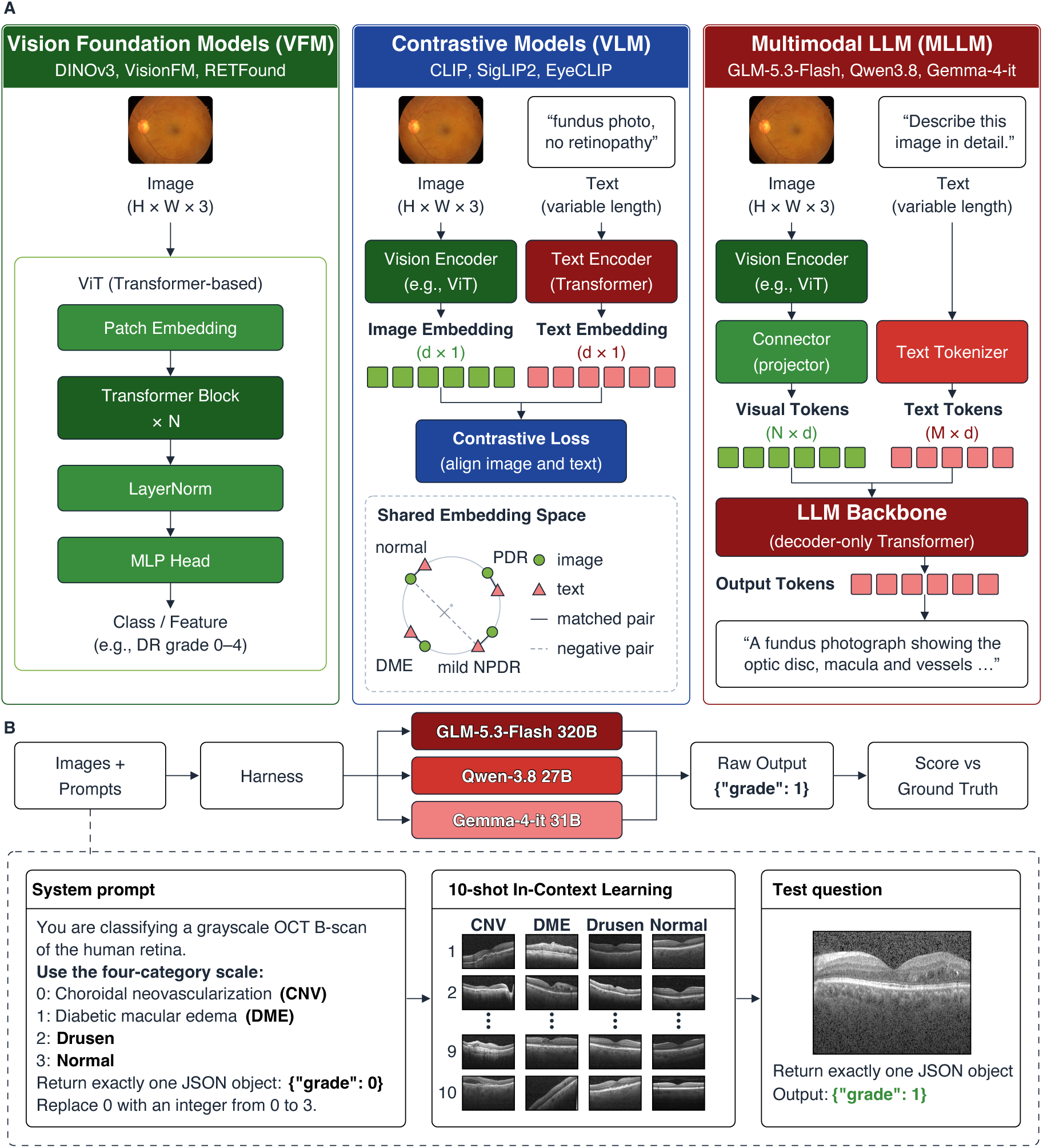
Model families and evaluation setup. **(A)** Vision models (e.g., DINOv3, RETFound, VisionFM) map an image to a label or feature vector. Contrastive VLMs (e.g., CLIP, SigLIP2, EyeClip) learn to encode and align images with text using a contrastive objective. MLLMs (e.g., GLM 5.3 Flash, Qwen 3.8, Gemma 4) project vision-encoder features into a decoder-only LLM that generates text. **(B)** Each image is sent with a fixed prompt to three MLLMs, whose JSON grades (0–3) are scored against ground truth. For few-shot ICL, labeled image examples are included in the prompt before the test image, without updating model parameters. The illustrated 10-shot setting includes 10 examples per class (40 total) across the four OCT categories (CNV, DME, drusen, and normal). Green, vision; red, language; blue, contrastive objective.

**Figure 2.**
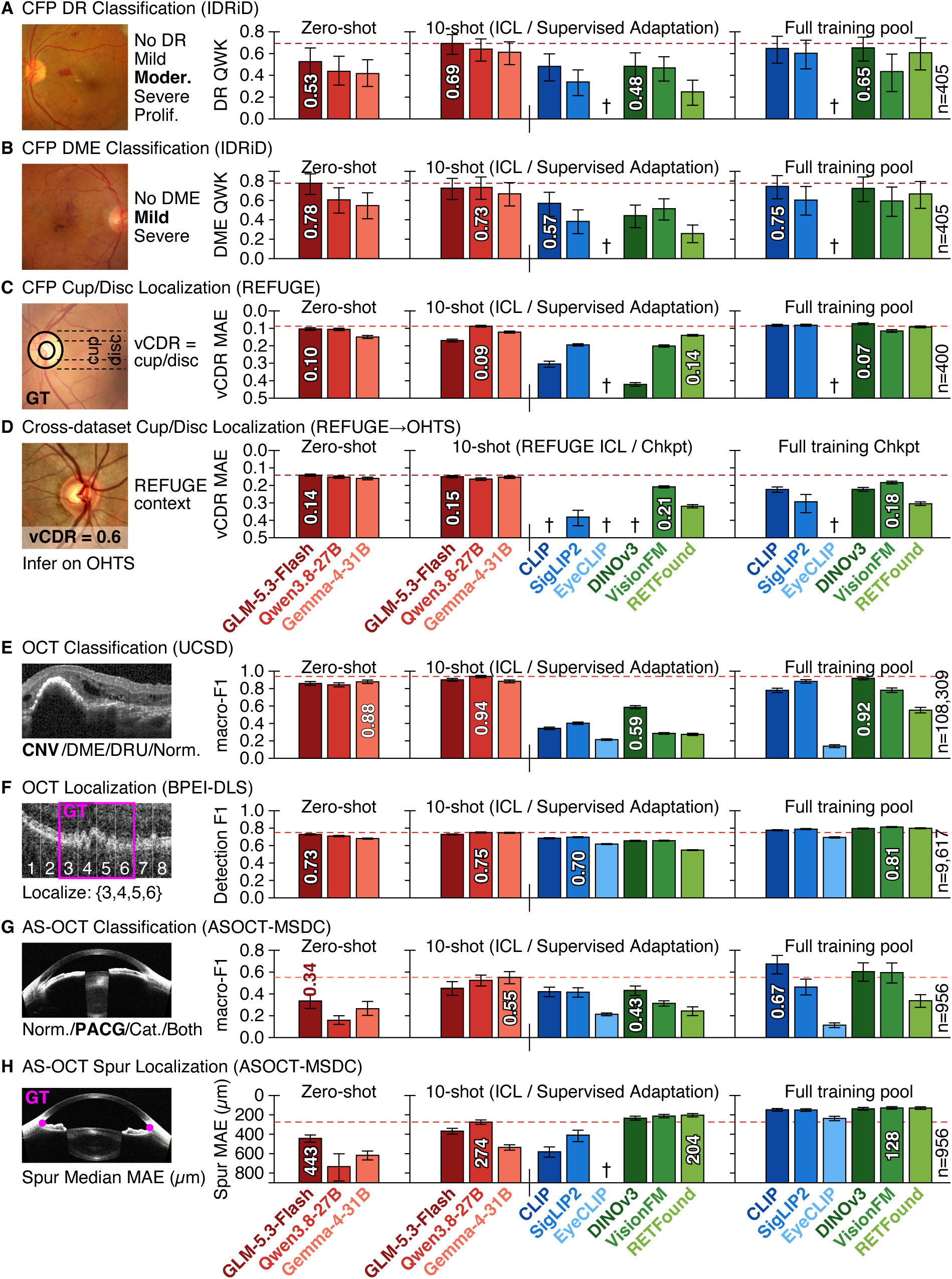
Quantitative comparison of ophthalmic classification and localization performance across model paradigms and clinical tasks. Classification and localization performance across model paradigms. For each task (A–H), a representative image with ground-truth (GT) annotation is shown at left; bars show zero-shot, 10-shot (in-context learning or supervised adaptation on identical images), and full-training-pool results. **(A)** CFP DR grading (IDRiD, QWK). **(B)** CFP DME grading (IDRiD, QWK). **(C)** Disc/cup localization with vCDR (REFUGE, MAE). **(D)** Cross-dataset vCDR transfer, REFUGE→OHTS (MAE). **(E)** OCT four-class classification (UCSD, macro-F1). **(F)** OCT band-level lesion localization (BPEI-DLS, detection F1). **(G)** AS-OCT angle classification (macro-F1). **(H)** AS-OCT scleral-spur localization (MAE). Red, MLLMs; blue, contrastive VLMs; green, VFMs.

**Figure 3.**
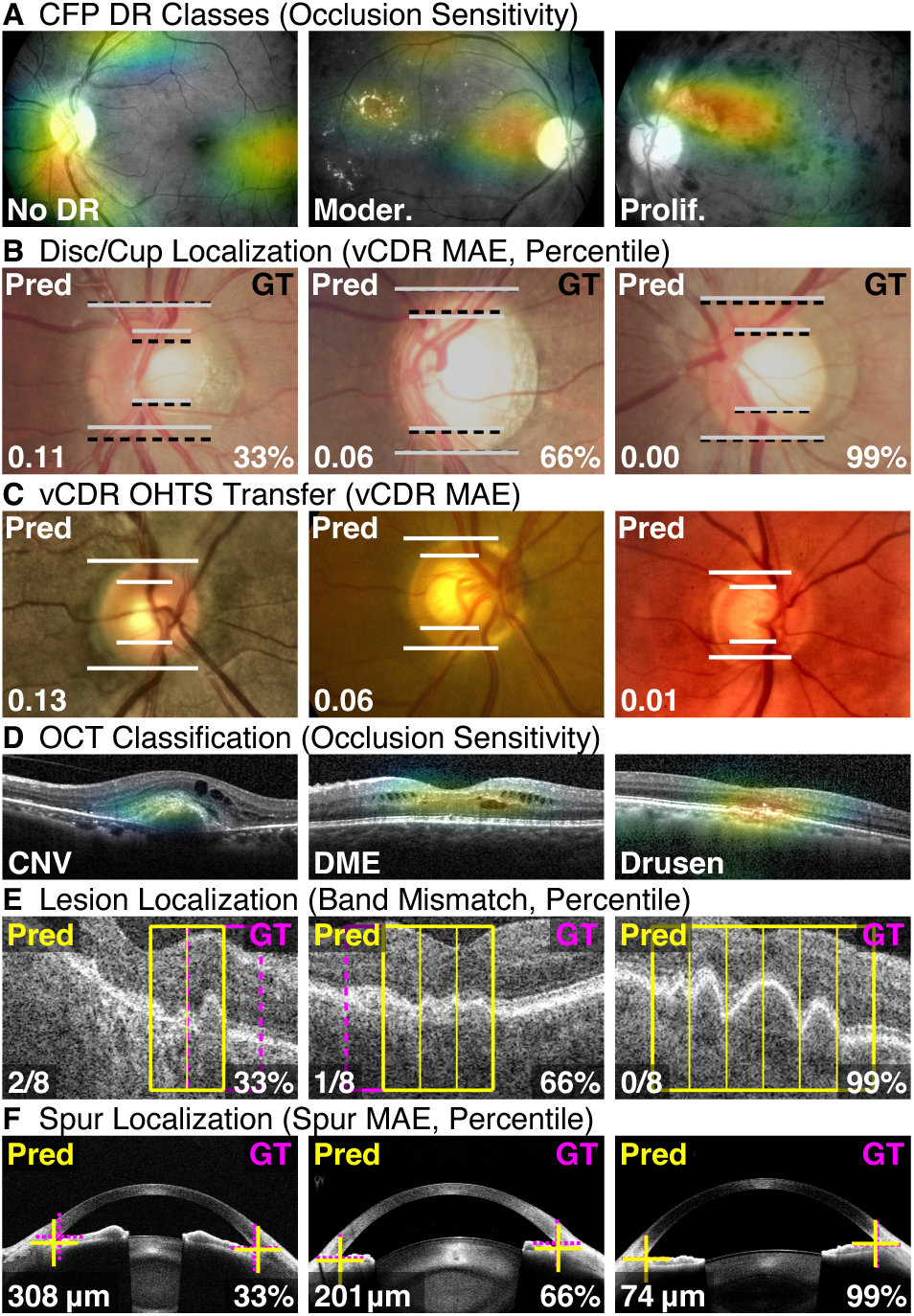
Representative MLLM localization predictions and visual grounding. **(A)** Occlusion sensitivity for DR severity classification on CFP: no DR, moderate, and proliferative DR. **(B)** Optic disc/cup localization on REFUGE with predicted vertical cup-to-disc ratio (vCDR); error and test-set percentile shown. **(C)** Cross-dataset vCDR estimation on OHTS using REFUGE-derived demonstrations. **(D)** Occlusion sensitivity for OCT disease classification (CNV, DME, drusen). **(E)** Band-level lesion localization on OCT: predicted versus ground-truth lesion bands. **(F)** Scleral-spur localization on AS-OCT: predicted versus reference landmark. In a and d, warm colors mark regions supporting the predicted class; occlusion maps reflect prediction dependence, not lesion segmentation. All rows use Qwen3.8-27B.

**Figure 4.**
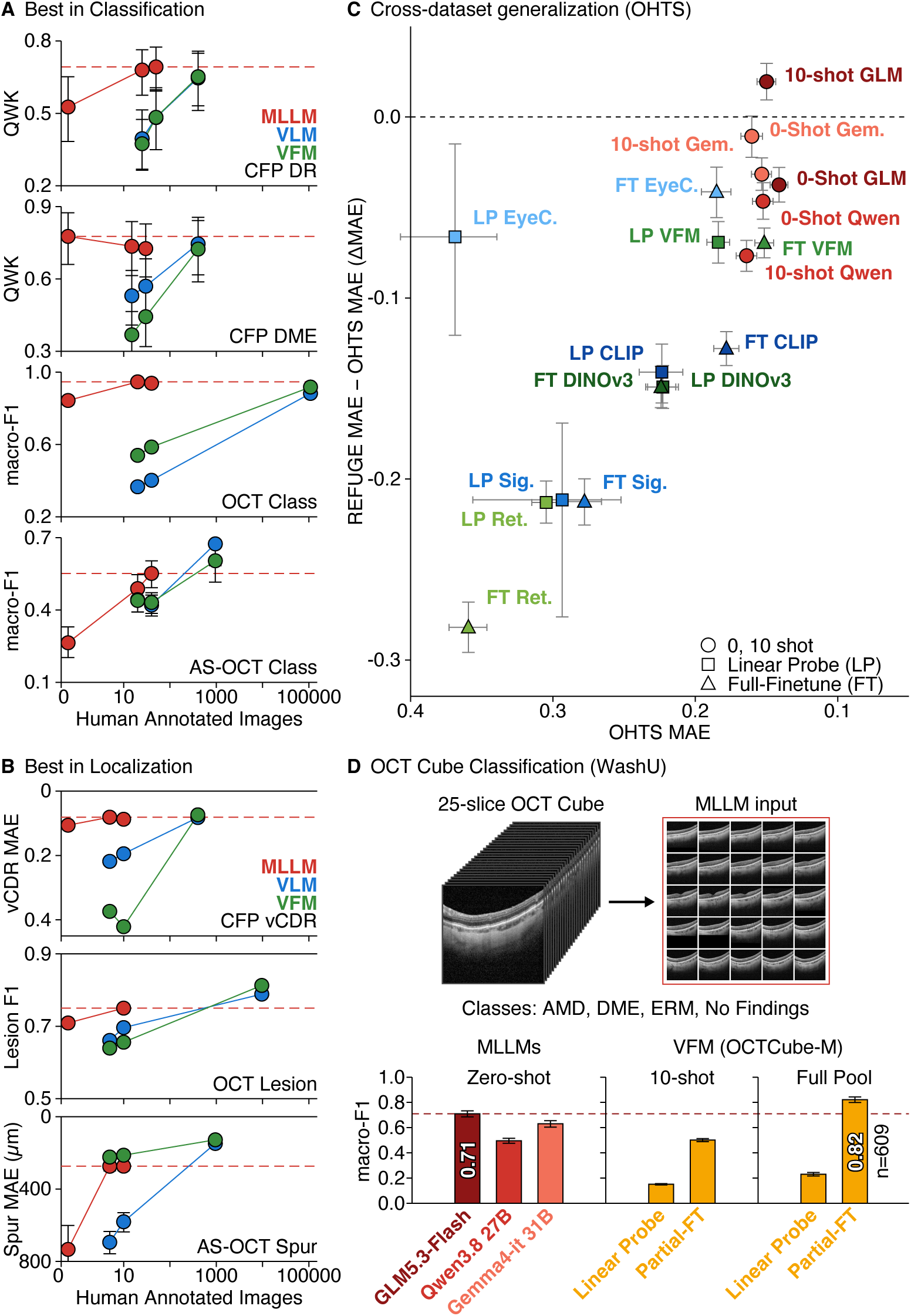
Label efficiency, cross-dataset generalization, and volumetric clinical OCT evaluation. (A, **B)** Best-in-class performance versus the number of labeled examples (zero-shot, 10-shot, full pool) for generative MLLMs (red), contrastive VLMs (blue), and VFMs (green) across classification/grading tasks **(A)** and localization tasks **(B)**. **(C)** Cross-dataset generalization from REFUGE to OHTS: OHTS vCDR MAE (horizontal, reversed; left is better) versus the change in MAE between datasets (vertical); zero-shot and 10-shot MLLMs (red circles) show the lowest OHTS error with the least degradation. **(D)** Zero-shot MLLM classification of private clinical OCT volumes (WashU; AMD, DME, ERM, no findings) compared with the volumetric foundation model OCTCube-M adapted on 10 volumes per class or the full development pool (n = 609). Error bars indicate bootstrap 95% CIs.

### DR Severity and DME Risk Grading

We evaluated five-grade DR severity and three-grade DME risk classification using IDRiD CFPs (Figure 2A–B), assessing recognition of retinal vascular abnormalities and macular hard exudates, respectively.

For DR grading (Figure 2A), GLM-5.3-Flash achieved the highest zero-shot quadratic-weighted kappa (QWK, 0.526; 95% CI, 0.384–0.652), compared with 0.437 for Qwen3.8-27B. With ten labeled examples per class (10-shot), GLM-5.3-Flash achieved the highest QWK (0.693; 95% CI, 0.592–0.774) using in-context learning without parameter updates, compared with 0.484 for DINOv3 and 0.482 for CLIP, the best VFM and contrastive VLM using linear probing, respectively. With full-data linear probing, DINOv3 achieved 0.652, improving to 0.714 with partial fine-tuning. SigLIP2 achieved 0.722 with full-data partial fine-tuning (Supplementary Appendix 1, A.13). Occlusion sensitivity analyses assessed clinically relevant image regions for both tasks (Figure 3a).

For DME grading (Figure 2B), GLM-5.3-Flash achieved the highest zero-shot QWK (0.776; 95% CI, 0.660–0.874). With 10-shot in-context learning, Qwen3.8-27B achieved the highest QWK (0.734; 95% CI, 0.611–0.841), compared with 0.514 for VisionFM and 0.570 for CLIP, the best VFM and contrastive VLM using linear probing, respectively. With full-data linear probing, DINOv3 achieved 0.724, improving to 0.745 with partial fine-tuning. SigLIP2 achieved 0.785 with full-data partial fine-tuning (Supplementary Appendix 1, B.13).

Overall, 10-shot MLLMs achieved substantial agreement with reference grades (QWK 0.69–0.73), approaching vision models adapted with substantially more labeled data (0.714–0.785). MLLM errors were predominantly adjacent-grade (adjacent accuracy 0.79–0.93), with high specificity, whereas supervised vision-model probes showed greater sensitivity for referable disease but more false positives among healthy eyes (Supplementary Appendix 1, A.13 and B.13).

### Glaucoma Classification and Cross-Dataset Generalization of vCDR

We evaluated vCDR estimation and glaucoma discrimination using REFUGE CFPs, followed by cross-dataset evaluation on OHTS.

For vCDR estimation (Figure 2C), GLM-5.3-Flash achieved the best zero-shot mean absolute error (MAE, 0.104; 95% CI, 0.096–0.111), compared with 0.106 (95% CI, 0.099–0.113) for Qwen3.8-27B. With 10-shot in-context learning, Qwen3.8-27B achieved the lowest MAE (0.087; 95% CI, 0.082–0.092), compared with 0.140 (95% CI, 0.135–0.145) for RETFound and 0.195 (95% CI, 0.189–0.201) for SigLIP2, the best VFM and contrastive VLM using linear probing, respectively. Demonstrations did not consistently improve performance: GLM-5.3-Flash worsened from 0.104 to 0.163–0.169, whereas Gemma-4-31B-it improved from 0.150 to 0.122. With full-data adaptation, DINOv3 achieved the lowest linear-probing MAE (0.074), while CLIP achieved the lowest overall MAE (0.050; 95% CI, 0.047–0.054) with full fine-tuning (Supplementary Appendix 1, C.1).

For glaucoma discrimination using predicted vCDR as a continuous screening score, zero-shot MLLMs achieved AUROCs of 0.810–0.828, highest for Qwen3.8-27B (0.828; 95% CI, 0.751–0.894). With 10-shot in-context learning, AUROCs ranged from 0.785 to 0.816, compared with 0.830 (95% CI, 0.783–0.872) for VisionFM and 0.679 (95% CI, 0.630–0.721) for CLIP using linear probing. Thus, zero-shot MLLMs approached the strongest 10-shot supervised probe without labeled examples. With full-data partial fine-tuning, RETFound achieved the highest AUROC (0.970; 95% CI, 0.936–0.993). VisionFM achieved the highest 10-shot probing AUROC despite higher vCDR MAE than RETFound (0.201 versus 0.140), indicating that measurement accuracy and discrimination were not directly aligned (Supplementary Appendix 1, C.2).

For cross-dataset evaluation, models were adapted using REFUGE examples and tested on OHTS without OHTS adaptation (Figure 2D). With 10-shot in-context learning, GLM-5.3-Flash achieved an MAE of 0.150 (95% CI, 0.144–0.157), compared with 0.164 (95% CI, 0.157–0.171) for Qwen3.8-27B. Among 10-shot linear probes, VisionFM achieved the lowest MAE (0.208; 95% CI, 0.202–0.214), followed by RETFound (0.320; 95% CI, 0.310–0.329) and SigLIP2 (0.382; 95% CI, 0.342–0.431). CLIP reached an MAE of 3.99, illustrating implausible unconstrained regression outputs under dataset shift. Under zero-shot evaluation, GLM-5.3-Flash achieved an MAE of 0.141 (95% CI, 0.135–0.147), compared with 0.152 for Qwen3.8-27B, outperforming both its 10-shot configuration (0.150) and the best 10-shot supervised probe (0.208) (Figure 4C).

Although zero-shot MLLMs achieved lower OHTS MAE (0.141–0.152) than the best 10-shot supervised probe (0.208), VisionFM showed a smaller change between REFUGE and OHTS (0.201 to 0.208) than 10-shot Qwen3.8-27B (0.087 to 0.164). Rankings also shifted: RETFound was the best 10-shot probe on REFUGE, whereas VisionFM performed best on OHTS. These findings indicate that task-specific demonstrations did not necessarily improve external-dataset performance.

To assess demographic robustness, vCDR estimation error was examined across OHTS subgroups stratified by age, sex, and race (Supplementary Appendix 1, D.2). MLLMs achieved lower MAE than 10-shot VisionFM across all subgroups. Across models, MAE was lower among Black than White participants (GLM zero-shot, 0.127 vs. 0.148) and among male than female participants (0.133 vs. 0.147), whereas no clear age-related trend was observed. Notably, the absolute Black–White MAE gap was 54.3% smaller for GLM zero-shot than for fully trained VisionFM (0.021 vs. 0.046), suggesting more consistent performance across racial subgroups. These exploratory findings support the cross-dataset generalizability of MLLMs, although the underlying causes of demographic differences warrant further investigation.

### OCT Macular Disease Classification

We evaluated four-class classification of normal retina, choroidal neovascularization (CNV), DME, and drusen using UCSD OCT (Figure 2E). Zero-shot MLLMs achieved macro-F1 scores of 0.843–0.877, highest for Gemma-4-31B-it (0.877; 95% CI, 0.857–0.897), compared with 0.100–0.156 for contrastive VLMs. With 10-shot in-context learning, Qwen3.8-27B achieved 0.938 (95% CI, 0.925–0.950), compared with 0.586 for DINOv3 and 0.402 for SigLIP2 using linear probing. With full-data linear probing, DINOv3 achieved 0.917 and SigLIP2 achieved 0.882. Full fine-tuning improved macro-F1 to 0.957 for VisionFM, 0.953 for RETFound, and 0.951 for DINOv3 (Supplementary Appendix 1, E.19). Occlusion sensitivity analyses assessed clinically relevant image regions (Figure 3d).

### BPEI-DLS: Retinal Pathology Localization

We evaluated localization of drusen and double-layer sign (DLS) using BPEI-DLS (Figure 2F). Each B-scan was divided into eight vertical bands, with models predicting pathology presence in each region. Vision models used supervised spatial prediction, whereas MLLMs selected numbered regions through Set-of-Marks (SoM) prompting.^46^

Under zero-shot evaluation, GLM-5.3-Flash achieved the highest MLLM lesion-level F1 (0.728; 95% CI, 0.722–0.735), compared with 0.709 for Qwen3.8-27B and 0.680 for Gemma-4-31B-it. With 10-shot in-context learning, Qwen3.8-27B achieved 0.750 (95% CI, 0.744–0.756), compared with 0.656 for VisionFM and 0.696 for SigLIP2 using linear probing. With full-data linear probing, VisionFM achieved 0.813, improving to 0.832 with partial fine-tuning. DINOv3 achieved the highest overall F1 (0.856; 95% CI, 0.851–0.861) with full-data partial fine-tuning (Supplementary Appendix 1, F.16). Representative predictions are shown in Figure 3e. Thus, MLLMs localized lesions competitively with limited supervision, whereas supervised vision models achieved higher performance with more labeled data.

### Anterior Chamber Angle Classification and Scleral-Spur Localization

We evaluated anterior-segment OCT (AS-OCT) interpretation through PACG and cataract classification and scleral-spur localization.

For multilabel classification of PACG and cataract, encompassing normal, cataract, PACG, and concurrent disease (Figure 2G), Gemma-4-31B-it achieved the highest zero-shot PACG macro-F1 (0.615; 95% CI, 0.535–0.691), compared with 0.583 for Qwen3.8-27B and 0.571 for GLM-5.3-Flash. With 10-shot in-context learning, Gemma-4-31B-it achieved 0.780 (95% CI, 0.721–0.832), compared with 0.670 for DINOv3 and 0.654 for SigLIP2 using linear probing. For additional tasks, Qwen3.8-27B achieved macro-F1 scores of 0.777 for cataract classification and 0.524 for four-class categorization with 10-shot in-context learning. With full-data partial fine-tuning, VisionFM achieved 0.912 for PACG and 0.804 for four-class classification (Supplementary Appendix 1, G.19).

For scleral-spur localization (Figure 2H), supervised vision models predicted Gaussian-smoothed spatial heatmaps, whereas MLLMs generated landmark coordinates from image instructions and marked few-shot demonstrations. Zero-shot GLM-5.3-Flash achieved the lowest MLLM localization error (442.7 µm; 95% CI, 407.3–481.3 µm). With 10-shot in-context learning, Qwen3.8-27B achieved 273.8 µm (95% CI, 251.9–297.8 µm), compared with 203.5 µm for RETFound and 408.7 µm for SigLIP2 using linear probing. With full-data linear probing, VisionFM achieved 127.9 µm, while SigLIP2 achieved 124.7 µm with full fine-tuning (Supplementary Appendix 1, H.1). Representative predictions are shown in Figure 3f. MLLMs performed competitively in classification, whereas fully adapted vision models achieved greater anatomical localization precision.

### Extension to Volumetric Clinical OCT Classification (WashU-OCT)

We evaluated four-class classification of healthy retina, AMD, DME, and ERM using 1,422 held-out EHR-linked WashU-OCT volumes.

For zero-shot MLLM evaluation, each volume was represented by 25 B-scans submitted together in a single multimodal inference request, enabling volume-level classification without parameter updates. GLM-5.3-Flash achieved accuracy of 0.730 (95% CI, 0.707–0.752) and macro-F1 of 0.710 (95% CI, 0.686–0.733), compared with 0.608 (95% CI, 0.584–0.634) and 0.496 (95% CI, 0.476–0.516), respectively, for Qwen3.8-27B, and 0.660 (95% CI, 0.635–0.684) and 0.631 (95% CI, 0.604–0.656), respectively, for Gemma-4-31B-it. GLM-5.3-Flash achieved class-specific F1 scores of 0.854 for AMD, 0.709 for DME, 0.691 for ERM, and 0.586 for healthy retina. Qwen3.8-27B performed substantially worse for ERM (F1, 0.039), frequently misclassifying ERM as healthy.

As a volumetric baseline, OCTCube-M, a three-dimensional Vision Transformer pretrained using masked autoencoding, processed volumes resized to 60 × 256 × 256 voxels. With ten labeled volumes per class, linear probing achieved accuracy of 0.316 (95% CI, 0.304–0.328) and macro-F1 of 0.151 (95% CI, 0.145–0.156) across five random seeds. Partial fine-tuning of the final two transformer blocks and classification head improved accuracy to 0.588 (95% CI, 0.573–0.602) and macro-F1 to 0.502 (95% CI, 0.489–0.513). With full-data linear probing, OCTCube-M achieved an accuracy of 0.390 (95% CI, 0.364–0.416) and a macro-F1 of 0.230 (95% CI, 0.216–0.244), improving to 0.854 (95% CI, 0.835–0.872) and 0.822 (95% CI, 0.799–0.843), respectively, with partial fine-tuning (Figure 4D).

Zero-shot GLM-5.3-Flash exceeded OCTCube-M under both 10-shot adaptation strategies (macro-F1, 0.710 versus 0.151 with linear probing and 0.502 with partial fine-tuning), although OCTCube-M performed better with full-data partial fine-tuning (0.822). These findings demonstrate the potential for training-free MLLM classification of clinical OCT volumes while highlighting the benefits of supervised adaptation for dedicated volumetric models.

## DISCUSSION

In this study, we built an evaluation harness to benchmark the performance of open-weight MLLMs compared to VFMs and VLMs for the interpretation of multiple ophthalmic image modalities encompassing a broad range of ocular conditions. The principal findings were first, that for OCT-based classification of retinal pathology as well as CFP-based severity and risk grading of DR and DME, MLLMs undergoing 10-shot in-context learning outperformed VFMs and VLMs trained on the same number of examples. Furthermore, MLLMs with 10 examples also approached the performance of VFMs and VLMs adapted using substantially larger labeled datasets on selected classification and grading tasks. Second, MLLMs demonstrated promising performance on institutionally curated datasets, including cross-dataset vCDR estimation and classification of clinical OCT volumes, outperforming the evaluated VFM and VLM baselines under limited-label conditions. Third, localization performance depended on task formulation: MLLMs outperformed vision models in region-level lesion localization under matched 10-shot conditions, whereas supervised vision models achieved greater precision in anatomical landmark localization.

This study demonstrates several advances that build upon prior benchmark studies assessing MLLM performance on ophthalmic image interpretation. Notably, the Foundation Ophthalmic Cross-Dataset Understanding under Shift (FOCUS) study previously characterized MLLMs on binary fundus classification across 10 public datasets,^19^ while the RetinaGroundBench study characterized zero-shot visual grounding of OCT biomarkers by open-weight MLLMs.^48^ Our present study expands on these foundations by evaluating MLLM performance using an in-context learning framework, in which models received clinical definitions and labeled examples matched across model families. In addition, while prior studies have evaluated MLLM performance with CFPs and individual OCT B-scans, we expand the repertoire of assessed imaging modalities to include AS-OCT and whole OCT volumes. Likewise, our study extends beyond previously studied tasks, such as binary classification and bounding-box grounding, to additional complex tasks, including severity grading, anatomical measurement, and disease status classification. Whereas prior studies have shown a limited ability of MLLMs to perform bounding-box grounding of OCT biomarkers without low-rank adaptation (LoRA) fine tuning,^48^ our work demonstrates that in-context learning allows MLLMs to perform region-level localization at a level similar to VFMs and VLMs trained on the same examples.

Altogether, our results mark a major progression in ophthalmic AI. Task-specific deep learning models have required training on thousands of graded two-dimensional images or annotated OCT volumes to achieve expert-level performance for a limited number of ophthalmic tasks.^1–6^ Moreover, despite a shift in much of this training burden to the pre-training phase for VFMs, our present study demonstrates that VFM performance still strongly depends on the amount of labeled training data provided for a given task.^11–14,17,18^ MLLMs offer a potential alternative to the heavy training requirements of other model architectures, since they can be directed to new tasks through prompting rather than training. Early evaluations of MLLMs in ophthalmology, however, found modest and variable diagnostic accuracy, with or without in-context examples.^24–29^ Our findings suggest that recent open-weight MLLMs, including Qwen3.8-27B, GLM-5.3-Flash, and Gemma-4-31B-it, can achieve substantially stronger performance on several ophthalmic imaging tasks than reported in earlier MLLM evaluations, particularly when provided with clinical definitions and a small number of labeled examples.

Several mechanisms may account for these results. Large-scale multimodal pretraining aligns visual features with language,^15,16^ which may enable MLLMs to apply written disease definitions to images. Prior work has linked zero-shot performance to the frequency of concepts in multimodal pretraining data.^49^ This may help explain why common, well-described findings such as macular edema, drusen and optic disc cupping were recognized without examples, although we did not test this mechanism directly. In-context examples may then align predictions with dataset-specific labeling conventions, allowing rapid adaptation to a new task without retraining or developing a specialized model. Task format may also help explain why MLLMs localized lesions well but not the scleral spur. Selecting among numbered regions, as in Set-of-Marks prompting,^46^ may reduce the spatial demands placed on MLLMs relative to generating precise coordinates as text.^48^ Together, these properties allow a single general-purpose MLLM to be applied to diverse ophthalmic tasks with only a few labeled examples.

Figure 4 summarizes the principal comparisons of the study. Panels A and B illustrate the contrasting relationships between labeled data availability and model performance: MLLMs achieved strong zero-shot and few-shot performance across classification tasks (Figure 4A) and, with a different profile, across localization tasks (Figure 4B), while supervised vision models generally benefited from larger training sets. Beyond label efficiency, zero-shot evaluation offers a different perspective on cross-dataset generalizability: rather than transferring a model adapted on one dataset to another, MLLMs can be evaluated directly on independent datasets without task-specific training. Consistent with this distinction, zero-shot MLLMs achieved lower vCDR estimation errors on OHTS than vision models adapted using REFUGE examples (Figure 4C). Panel D extends this comparison to volumetric data: the strongest zero-shot MLLM classification of private clinical OCT volumes exceeded a volumetric vision foundation model adapted on ten examples per class.

Training-data contamination is a central concern in benchmarking MLLMs. Evidence of contamination with widely used multimodal benchmarks has been reported in both open-weight and proprietary MLLMs.^50^ Public ophthalmic datasets are additionally susceptible to leakage between training and test splits.^51^ Public datasets remain essential for reproducibility and comparison across studies, and recent benchmarks, as well as our own, have relied on them.^19,48^ To address this issue, we utilized three non-publicly released datasets to evaluate model generalizability, which include the controlled access OHTS as well as the institutionally curated BPEI-DLS and WashU-OCT datasets. On all three private datasets, MLLMs outperformed VFMs and VLMs at a matched label budget. Furthermore, for OHTS, BPEI-DLS, and WashU-OCT, zero-shot MLLMs outperformed VFMs and VLMs trained on 10 labeled examples per class. Therefore, the consistent performance between private and public datasets strengthens confidence that our public benchmark results reflect generalization rather than memorization.

Generalization also depends on whether performance holds across patient populations. In this study, the 10 labeled examples per class were sampled randomly, without stratification by age, sex, race, or ethnicity, so a small example set could in principle limit generalizability. However, zero-shot inference uses no examples, and as noted above, zero-shot MLLMs achieved the lowest vCDR error on OHTS, a multicamera US cohort in which 26% of participants were Black.^39^ Moreover, 10-shot demonstrations drawn from REFUGE, a dataset of Chinese patients,^38^ increased OHTS error only slightly relative to zero-shot inference (MAE, 0.150 vs 0.141). Across race, sex, and age subgroups on OHTS, MLLMs achieved lower error than vision models adapted with the same 10 examples, and no subgroup’s error exceeded the overall error by more than 0.02 (Supplementary Appendix 1, D.2). Error was modestly lower among Black and male participants for all models, including the fully trained vision model, suggesting differences in image or optic disc characteristics between subgroups rather than model-specific bias. Nevertheless, these analyses reflect performance on a single task, and because the demographic composition of MLLM pretraining data is undisclosed, selecting representative examples and validating models on target populations continues to remain important before clinical use.

This study has several limitations. First, although the benchmark spanned multiple tasks, imaging modalities, and ocular diseases, it did not encompass all ophthalmic tasks. Second, all tasks in this study involved findings that clinicians can recognize. Fundus photographs also contain systemic information not readily quantified through routine clinical assessment. In the Atherosclerosis Risk in Communities Study, standardized grading showed that retinal arteriolar narrowing was associated with current and past blood pressure,^52,53^ and deep learning has since been used to predict cardiovascular risk factors from fundus photographs.^54^ Whether in-context learning can identify such signals was not tested here. Third, we did not assess semantic segmentation. MLLMs could represent segmentation masks as text, such as binary masks, but this approach has not been fully evaluated, so it remains uncertain whether dedicated segmentation models will remain better suited for this task. Fourth, we adopted standardized evaluation protocols, using primarily linear probing with partial fine-tuning for selected arms, for few-shot VFM and VLM adaptation and in-context learning without parameter updates for MLLMs. Although alternative fine-tuning strategies and prompt designs may influence performance, our objective was to compare model families under a matched label budget and consistent evaluation settings rather than optimize each model individually. Finally, the field is evolving rapidly, and our results represent a single point in time. With continued investment in general-purpose models, the advantage of MLLMs in label-limited settings may widen, although this will require reassessment as models and methods evolve. Our evaluation harness, including task prompts, labeled example sets, and inference settings, will be released as open-source software (https://github.com/wusmai/Ophthalmic-AI-with-Ten-Examples) to facilitate further development and evaluation of MLLMs for ophthalmic image analysis.

Our work suggests several directions for future research. Parameter-efficient fine-tuning methods such as LoRA may improve MLLM performance beyond in-context learning.^55^ LoRA fine-tuning improved OCT biomarker grounding in RetinaGroundBench,^48^ and open-weight MLLMs can be fine-tuned on local hardware.^21^ Hybrid systems that combine MLLMs with dedicated segmentation models may also improve localization precision. Because open-weight models can be deployed within institutional infrastructure, they facilitate privacy-preserving clinical research, although HIPAA requirements continue to apply.^30–32^ Broader external and prospective validation will be essential. In a randomized controlled trial, an ophthalmic foundation model used as a clinical assistant improved diagnostic and referral accuracy,^56^ and comparable evidence will be required before MLLM-based tools enter clinical practice.

Developing task-specific ophthalmic AI has often required substantial annotation effort, computational resources, machine learning expertise, and a dedicated model for each application, which can limit development to centers with those resources. Because locally deployed MLLMs can be applied to new tasks using task-specific instructions and a small number of labeled examples, effort could shift from annotation and model training toward defining clinically meaningful questions, curating representative examples, and rigorously evaluating model outputs. This shift could democratize ophthalmic AI, allowing clinicians to develop and validate tools for the questions most relevant to their own patients, including rare diseases with few labeled cases and underserved populations, while keeping patient data within the institution.

In conclusion, locally deployed, open-weight MLLMs with 10 labeled examples per class outperformed VFMs and VLMs trained on the same examples in most of the ophthalmic tasks evaluated and generalized to new datasets without retraining. These findings highlight a broader opportunity: making advanced ophthalmic image analysis accessible without massive labeled datasets or specialized model development. If validated prospectively, these findings suggest that the next phase of ophthalmic AI will be defined less by the size of labeled datasets and more by how effectively general-purpose MLLMs are directed toward clinical needs.

## Declaration of Generative AI and AI-assisted technologies in the writing process

During the preparation of this work the authors used Claude and a locally hosted instance of GLM-5.3-Flash to assist with literature review, draft an initial manuscript outline, correct grammatical errors, and improve the clarity and readability of the text. After using these tools, the authors reviewed and edited the content as needed and take full responsibility for the integrity and accuracy of the publication.

## Data Availability

The public datasets analyzed in this study (UCSD OCT, IDRiD, REFUGE, and ASOCT-MSDC) are available from their original sources as cited in the manuscript. OHTS data are available from the Ocular Hypertension Treatment Study Group upon application and approval. The BPEI-DLS and WashU-OCT datasets are not publicly available because they contain protected patient information. Task prompts, labeled example sets, and evaluation code will be available at https://github.com/wusmai/Ophthalmic-AI-with-Ten-Examples.

