## Appendix 1 for "Ophthalmic AI With Just Ten Examples: Comparing Multimodal Large Language Models, Vision Foundation Models, and Vision-Language Models"

Estimation protocol: every metric uses an image-level paired bootstrap (10,000 percentile replicates, seed 2026); invalid predictions are excluded per arm. Cells are point estimates with the 95% CI in parentheses. Confusion matrices pool counts over training seeds. Class-wise precision/recall/F1 are pooled point estimates.

**Figure 2, Panel A - CFP DR Classification (IDRiD)**

Dataset size: train+val n = 405 images; test n = 102 images.

Nine of the 516 official IDRiD images are excluded in every arm (test: IDRiD_064; train: IDRiD_021, _028, _045, _279, _327, _333, _375, _391), identified as duplicates by image hashing in the original grading pipeline; the exclusion is an explicit identifier set applied identically to training and evaluation.

**A.1 GLM zero-shot**

| True \ Pred | grade 0 | grade 1 | grade 2 | grade 3 | grade 4 | Precision | Recall | F1 |
|---|---|---|---|---|---|---|---|---|
| grade 0 | 18 | 16 | 0 | 0 | 0 | 0.621 | 0.529 | 0.571 |
| grade 1 | 2 | 3 | 0 | 0 | 0 | 0.088 | 0.600 | 0.154 |
| grade 2 | 7 | 8 | 17 | 0 | 0 | 0.500 | 0.531 | 0.515 |
| grade 3 | 1 | 4 | 12 | 1 | 0 | 0.500 | 0.056 | 0.100 |
| grade 4 | 1 | 3 | 5 | 1 | 3 | 1.000 | 0.231 | 0.375 |

**A.2 Qwen zero-shot**

| True \ Pred | grade 0 | grade 1 | grade 2 | grade 3 | grade 4 | Precision | Recall | F1 |
|---|---|---|---|---|---|---|---|---|
| grade 0 | 34 | 0 | 0 | 0 | 0 | 0.586 | 1.000 | 0.739 |
| grade 1 | 5 | 0 | 0 | 0 | 0 | 0.000 | 0.000 | 0.000 |
| grade 2 | 13 | 5 | 12 | 2 | 0 | 0.545 | 0.375 | 0.444 |
| grade 3 | 3 | 4 | 9 | 2 | 0 | 0.333 | 0.111 | 0.167 |
| grade 4 | 3 | 6 | 1 | 2 | 1 | 1.000 | 0.077 | 0.143 |

**A.3 Gemma zero-shot**

| True \ Pred | grade 0 | grade 1 | grade 2 | grade 3 | grade 4 | Precision | Recall | F1 |
|---|---|---|---|---|---|---|---|---|
| grade 0 | 34 | 0 | 0 | 0 | 0 | 0.557 | 1.000 | 0.716 |
| grade 1 | 5 | 0 | 0 | 0 | 0 | 0.000 | 0.000 | 0.000 |
| grade 2 | 15 | 3 | 14 | 0 | 0 | 0.438 | 0.438 | 0.438 |
| grade 3 | 3 | 2 | 13 | 0 | 0 | 0.000 | 0.000 | 0.000 |
| grade 4 | 4 | 3 | 5 | 1 | 0 | 0.000 | 0.000 | 0.000 |

**A.4 GLM 10-shot ICL**

| True \ Pred | grade 0 | grade 1 | grade 2 | grade 3 | grade 4 | Precision | Recall | F1 |
|---|---|---|---|---|---|---|---|---|
| grade 0 | 98 | 57 | 15 | 0 | 0 | 0.742 | 0.576 | 0.649 |
| grade 1 | 12 | 10 | 3 | 0 | 0 | 0.085 | 0.400 | 0.141 |
| grade 2 | 17 | 41 | 57 | 45 | 0 | 0.432 | 0.356 | 0.390 |
| grade 3 | 5 | 5 | 29 | 39 | 12 | 0.402 | 0.433 | 0.417 |
| grade 4 | 0 | 4 | 28 | 13 | 20 | 0.625 | 0.308 | 0.412 |

**A.5 Qwen 10-shot ICL**

| True \ Pred | grade 0 | grade 1 | grade 2 | grade 3 | grade 4 | Precision | Recall | F1 |
|---|---|---|---|---|---|---|---|---|
| grade 0 | 125 | 38 | 5 | 0 | 2 | 0.665 | 0.735 | 0.698 |
| grade 1 | 20 | 5 | 0 | 0 | 0 | 0.061 | 0.200 | 0.093 |
| grade 2 | 35 | 25 | 52 | 32 | 16 | 0.433 | 0.325 | 0.371 |

| | | | | | | | | |
|---|---|---|---|---|---|---|---|---|
| grade 3 | 6 | 6 | 34 | 35 | 9 | 0.493 | 0.389 | 0.435 |
| grade 4 | 2 | 8 | 29 | 4 | 22 | 0.449 | 0.338 | 0.386 |

**A.6 Gemma 10-shot ICL**

| True \ Pred | grade 0 | grade 1 | grade 2 | grade 3 | grade 4 | Precision | Recall | F1 |
|---|---|---|---|---|---|---|---|---|
| grade 0 | 125 | 35 | 7 | 1 | 2 | 0.616 | 0.735 | 0.670 |
| grade 1 | 23 | 2 | 0 | 0 | 0 | 0.031 | 0.080 | 0.044 |
| grade 2 | 46 | 16 | 60 | 30 | 8 | 0.420 | 0.375 | 0.396 |
| grade 3 | 7 | 8 | 39 | 33 | 3 | 0.478 | 0.367 | 0.415 |
| grade 4 | 2 | 4 | 37 | 5 | 17 | 0.567 | 0.262 | 0.358 |

**A.7 CLIP linear-probe, 10 shot**

| True \ Pred | grade 0 | grade 1 | grade 2 | grade 3 | grade 4 | Precision | Recall | F1 |
|---|---|---|---|---|---|---|---|---|
| grade 0 | 68 | 53 | 19 | 28 | 2 | 0.642 | 0.400 | 0.493 |
| grade 1 | 9 | 6 | 5 | 5 | 0 | 0.079 | 0.240 | 0.119 |
| grade 2 | 15 | 11 | 40 | 72 | 22 | 0.440 | 0.250 | 0.319 |
| grade 3 | 5 | 3 | 13 | 52 | 17 | 0.287 | 0.578 | 0.384 |
| grade 4 | 9 | 3 | 14 | 24 | 15 | 0.268 | 0.231 | 0.248 |

**A.8 SigLIP2 linear-probe, 10 shot**

| True \ Pred | grade 0 | grade 1 | grade 2 | grade 3 | grade 4 | Precision | Recall | F1 |
|---|---|---|---|---|---|---|---|---|
| grade 0 | 25 | 107 | 8 | 19 | 11 | 0.431 | 0.147 | 0.219 |
| grade 1 | 4 | 16 | 0 | 4 | 1 | 0.073 | 0.640 | 0.131 |
| grade 2 | 18 | 50 | 19 | 45 | 28 | 0.487 | 0.119 | 0.191 |
| grade 3 | 6 | 28 | 6 | 33 | 17 | 0.292 | 0.367 | 0.325 |
| grade 4 | 5 | 18 | 6 | 12 | 24 | 0.296 | 0.369 | 0.329 |

**A.9 EyeCLIP linear-probe, 10 shot**

| True \ Pred | grade 0 | grade 1 | grade 2 | grade 3 | grade 4 | Precision | Recall | F1 |
|---|---|---|---|---|---|---|---|---|
| grade 0 | 37 | 65 | 10 | 15 | 43 | 0.394 | 0.218 | 0.280 |
| grade 1 | 5 | 7 | 3 | 2 | 8 | 0.036 | 0.280 | 0.064 |
| grade 2 | 29 | 57 | 14 | 9 | 51 | 0.368 | 0.087 | 0.141 |
| grade 3 | 9 | 42 | 5 | 11 | 23 | 0.282 | 0.122 | 0.171 |
| grade 4 | 14 | 22 | 6 | 2 | 21 | 0.144 | 0.323 | 0.199 |

**A.10 DINOv3 linear-probe, 10 shot**

| True \ Pred | grade 0 | grade 1 | grade 2 | grade 3 | grade 4 | Precision | Recall | F1 |
|---|---|---|---|---|---|---|---|---|
| grade 0 | 27 | 87 | 19 | 23 | 14 | 0.711 | 0.159 | 0.260 |
| grade 1 | 2 | 16 | 5 | 2 | 0 | 0.113 | 0.640 | 0.192 |
| grade 2 | 6 | 35 | 30 | 60 | 29 | 0.385 | 0.188 | 0.252 |
| grade 3 | 1 | 1 | 11 | 41 | 36 | 0.283 | 0.456 | 0.349 |
| grade 4 | 2 | 3 | 13 | 19 | 28 | 0.262 | 0.431 | 0.326 |

**A.11 VisionFM linear-probe, 10 shot**

| True \ Pred | grade 0 | grade 1 | grade 2 | grade 3 | grade 4 | Precision | Recall | F1 |
|---|---|---|---|---|---|---|---|---|
| grade 0 | 72 | 31 | 35 | 29 | 3 | 0.600 | 0.424 | 0.497 |
| grade 1 | 14 | 6 | 2 | 3 | 0 | 0.109 | 0.240 | 0.150 |

| | | | | | | | | |
|---|---|---|---|---|---|---|---|---|
| grade 2 | 24 | 16 | 48 | 52 | 20 | 0.390 | 0.300 | 0.339 |
| grade 3 | 2 | 1 | 20 | 45 | 22 | 0.296 | 0.500 | 0.372 |
| grade 4 | 8 | 1 | 18 | 23 | 15 | 0.250 | 0.231 | 0.240 |

### A.12 RETFound linear-probe, 10 shot

| True \ Pred | grade 0 | grade 1 | grade 2 | grade 3 | grade 4 | Precision | Recall | F1 |
|---|---|---|---|---|---|---|---|---|
| grade 0 | 30 | 42 | 37 | 41 | 20 | 0.500 | 0.176 | 0.261 |
| grade 1 | 3 | 4 | 7 | 8 | 3 | 0.049 | 0.160 | 0.075 |
| grade 2 | 16 | 20 | 34 | 60 | 30 | 0.370 | 0.212 | 0.270 |
| grade 3 | 2 | 12 | 5 | 46 | 25 | 0.263 | 0.511 | 0.347 |
| grade 4 | 9 | 4 | 9 | 20 | 23 | 0.228 | 0.354 | 0.277 |

### A.13 Bootstrapped metrics, IDRiD DR (all bootstrapped metrics)

| Arm | accuracy | macro f1 | qwk | balanced accuracy |
|---|---|---|---|---|
| GLM zero-shot | 0.412 (0.314-0.510) | 0.343 (0.246-0.435) | 0.526 (0.384-0.652) | 0.389 (0.264-0.507) |
| Qwen zero-shot | 0.480 (0.382-0.578) | 0.299 (0.221-0.379) | 0.437 (0.311-0.575) | 0.313 (0.263-0.373) |
| Gemma zero-shot | 0.471 (0.373-0.569) | 0.231 (0.188-0.270) | 0.416 (0.298-0.543) | 0.287 (0.253-0.325) |
| CLIP zero-shot | 0.176 (0.108-0.255) | 0.060 (0.039-0.081) | 0.000 (0.000-0.000) | 0.200 (0.200-0.200) |
| SigLIP2 zero-shot | 0.147 (0.078-0.216) | 0.103 (0.040-0.164) | 0.089 (0.020-0.169) | 0.280 (0.200-0.400) |
| EyeCLIP zero-shot | 0.333 (0.245-0.422) | 0.100 (0.079-0.119) | 0.000 (0.000-0.000) | 0.200 (0.200-0.200) |
| GLM 5-shot ICL | 0.463 (0.376-0.547) | 0.414 (0.326-0.492) | 0.680 (0.575-0.764) | 0.434 (0.331-0.538) |
| Qwen 5-shot ICL | 0.494 (0.406-0.582) | 0.387 (0.307-0.455) | 0.652 (0.539-0.747) | 0.373 (0.303-0.446) |
| Gemma 5-shot ICL | 0.490 (0.406-0.575) | 0.359 (0.283-0.425) | 0.579 (0.453-0.690) | 0.352 (0.292-0.418) |
| CLIP linear-probe, 5 shot | 0.345 (0.296-0.394) | 0.319 (0.267-0.365) | 0.395 (0.265-0.514) | 0.357 (0.307-0.407) |
| CLIP partial-FT (last-2 layers), 5 shot | 0.355 (0.306-0.404) | 0.321 (0.268-0.366) | 0.439 (0.309-0.552) | 0.344 (0.283-0.407) |
| SigLIP2 linear-probe, 5 shot | 0.253 (0.212-0.292) | 0.232 (0.187-0.271) | 0.212 (0.120-0.300) | 0.284 (0.246-0.323) |
| SigLIP2 partial-FT (last-2 layers), 5 shot | 0.245 (0.194-0.298) | 0.238 (0.183-0.288) | 0.388 (0.284-0.481) | 0.317 (0.259-0.377) |
| EyeCLIP linear-probe, 5 shot | 0.200 (0.165-0.237) | 0.171 (0.141-0.197) | 0.009 (-0.011-0.033) | 0.206 (0.176-0.237) |
| EyeCLIP partial-FT (last-2 layers), 5 shot | 0.204 (0.171-0.237) | 0.183 (0.154-0.208) | -0.026 (-0.060-0.009) | 0.217 (0.187-0.244) |
| DINOv3 linear-probe, 5 shot | 0.290 (0.233-0.349) | 0.281 (0.211-0.343) | 0.374 (0.269-0.475) | 0.336 (0.247-0.439) |
| DINOv3 partial-FT (last-2 layers), 5 shot | 0.296 (0.241-0.355) | 0.289 (0.224-0.346) | 0.405 (0.299-0.500) | 0.341 (0.265-0.424) |
| VisionFM linear-probe, 5 shot | 0.302 (0.257-0.349) | 0.271 (0.226-0.310) | 0.342 (0.230-0.444) | 0.293 (0.254-0.335) |
| VisionFM partial-FT (last-2 layers), 5 shot | 0.296 (0.253-0.343) | 0.273 (0.227-0.315) | 0.362 (0.261-0.453) | 0.320 (0.275-0.368) |

| | | | | |
|---|---|---|---|---|
| RETFound linear-probe, 5 shot | 0.282 (0.225-0.343) | 0.226 (0.179-0.269) | 0.137 (0.035-0.237) | 0.266 (0.221-0.313) |
| RETFound partial-FT (last-2 layers), 5 shot | 0.294 (0.235-0.355) | 0.246 (0.194-0.293) | 0.164 (0.065-0.262) | 0.283 (0.233-0.337) |
| GLM 10-shot ICL | 0.439 (0.355-0.525) | 0.402 (0.313-0.483) | 0.693 (0.592-0.774) | 0.415 (0.310-0.526) |
| Qwen 10-shot ICL | 0.469 (0.390-0.549) | 0.397 (0.319-0.465) | 0.641 (0.530-0.734) | 0.398 (0.317-0.486) |
| Gemma 10-shot ICL | 0.465 (0.382-0.545) | 0.377 (0.293-0.448) | 0.612 (0.498-0.708) | 0.364 (0.290-0.444) |
| CLIP linear-probe, 10 shot | 0.355 (0.298-0.414) | 0.312 (0.255-0.365) | 0.482 (0.349-0.598) | 0.340 (0.280-0.406) |
| CLIP partial-FT (last-2 layers), 10 shot | 0.390 (0.333-0.449) | 0.346 (0.284-0.404) | 0.568 (0.457-0.659) | 0.364 (0.291-0.456) |
| SigLIP2 linear-probe, 10 shot | 0.229 (0.180-0.280) | 0.239 (0.186-0.286) | 0.339 (0.214-0.450) | 0.328 (0.254-0.399) |
| SigLIP2 partial-FT (last-2 layers), 10 shot | 0.275 (0.216-0.335) | 0.275 (0.212-0.333) | 0.474 (0.349-0.587) | 0.384 (0.308-0.452) |
| EyeCLIP linear-probe, 10 shot | 0.176 (0.151-0.204) | 0.171 (0.144-0.194) | 0.041 (-0.001-0.081) | 0.206 (0.178-0.234) |
| EyeCLIP partial-FT (last-2 layers), 10 shot | 0.171 (0.143-0.198) | 0.165 (0.138-0.188) | 0.011 (-0.038-0.060) | 0.185 (0.137-0.243) |
| DINOv3 linear-probe, 10 shot | 0.278 (0.225-0.335) | 0.276 (0.217-0.329) | 0.484 (0.349-0.606) | 0.375 (0.310-0.443) |
| DINOv3 partial-FT (last-2 layers), 10 shot | 0.290 (0.237-0.347) | 0.288 (0.227-0.343) | 0.527 (0.415-0.624) | 0.376 (0.310-0.445) |
| VisionFM linear-probe, 10 shot | 0.365 (0.308-0.420) | 0.320 (0.263-0.366) | 0.468 (0.348-0.571) | 0.339 (0.278-0.395) |
| VisionFM partial-FT (last-2 layers), 10 shot | 0.320 (0.269-0.373) | 0.299 (0.242-0.351) | 0.432 (0.307-0.542) | 0.352 (0.283-0.424) |
| RETFound linear-probe, 10 shot | 0.269 (0.210-0.329) | 0.246 (0.190-0.298) | 0.249 (0.126-0.356) | 0.283 (0.219-0.348) |
| RETFound partial-FT (last-2 layers), 10 shot | 0.294 (0.229-0.361) | 0.261 (0.203-0.314) | 0.280 (0.153-0.394) | 0.285 (0.223-0.350) |
| CLIP linear-probe, full dataset | 0.578 (0.480-0.676) | 0.428 (0.341-0.508) | 0.647 (0.512-0.758) | 0.430 (0.352-0.515) |
| CLIP partial-FT (last-2 layers), full dataset | 0.500 (0.402-0.598) | 0.377 (0.290-0.454) | 0.543 (0.387-0.672) | 0.381 (0.299-0.468) |
| SigLIP2 linear-probe, full dataset | 0.539 (0.441-0.637) | 0.407 (0.310-0.488) | 0.604 (0.459-0.723) | 0.400 (0.319-0.484) |
| SigLIP2 partial-FT (last-2 layers), full dataset | 0.559 (0.461-0.657) | 0.429 (0.333-0.511) | 0.722 (0.619-0.807) | 0.436 (0.353-0.522) |
| EyeCLIP linear-probe, full dataset | 0.402 (0.304-0.500) | 0.195 (0.153-0.234) | 0.069 (-0.089-0.237) | 0.249 (0.201-0.298) |
| EyeCLIP partial-FT (last-2 layers), full dataset | 0.402 (0.304-0.500) | 0.197 (0.153-0.235) | 0.191 (0.012-0.363) | 0.250 (0.204-0.296) |
| DINOv3 linear-probe, full dataset | 0.500 (0.402-0.598) | 0.394 (0.303-0.473) | 0.652 (0.532-0.750) | 0.395 (0.308-0.485) |
| DINOv3 partial-FT (last-2 layers), full dataset | 0.608 (0.510-0.696) | 0.472 (0.380-0.551) | 0.714 (0.605-0.803) | 0.472 (0.389-0.559) |
| VisionFM linear-probe, full dataset | 0.471 (0.373-0.569) | 0.359 (0.266-0.443) | 0.435 (0.251-0.596) | 0.366 (0.287-0.452) |
| VisionFM partial-FT (last-2 layers), full dataset | 0.549 (0.451-0.647) | 0.435 (0.341-0.517) | 0.569 (0.396-0.711) | 0.435 (0.350-0.524) |

| | | | | |
|---|---|---|---|---|
| RETFound linear-probe, full dataset | 0.559 (0.461-0.657) | 0.421 (0.325-0.502) | 0.608 (0.450-0.743) | 0.440 (0.361-0.522) |
| RETFound partial-FT (last-2 layers), full dataset | 0.500 (0.402-0.598) | 0.399 (0.307-0.480) | 0.522 (0.363-0.660) | 0.396 (0.313-0.484) |

**Figure 2, Panel B - CFP DME Classification (IDRiD)**

Dataset size: train+val n = 405 images; test n = 102 images.

Nine of the 516 official IDRiD images are excluded in every arm (test: IDRiD_064; train: IDRiD_021, _028, _045, _279, _327, _333, _375, _391), identified as duplicates by image hashing in the original grading pipeline; the exclusion is an explicit identifier set applied identically to training and evaluation.

**B.1 GLM zero-shot**

| True \ Pred | grade 0 | grade 1 | grade 2 | Precision | Recall | F1 |
|---|---|---|---|---|---|---|
| grade 0 | 43 | 1 | 0 | 0.741 | 0.977 | 0.843 |
| grade 1 | 8 | 2 | 0 | 0.250 | 0.200 | 0.222 |
| grade 2 | 7 | 5 | 36 | 1.000 | 0.750 | 0.857 |

**B.2 Qwen zero-shot**

| True \ Pred | grade 0 | grade 1 | grade 2 | Precision | Recall | F1 |
|---|---|---|---|---|---|---|
| grade 0 | 42 | 1 | 1 | 0.689 | 0.955 | 0.800 |
| grade 1 | 8 | 2 | 0 | 0.118 | 0.200 | 0.148 |
| grade 2 | 11 | 14 | 23 | 0.958 | 0.479 | 0.639 |

**B.3 Gemma zero-shot**

| True \ Pred | grade 0 | grade 1 | grade 2 | Precision | Recall | F1 |
|---|---|---|---|---|---|---|
| grade 0 | 42 | 2 | 0 | 0.656 | 0.955 | 0.778 |
| grade 1 | 8 | 2 | 0 | 0.105 | 0.200 | 0.138 |
| grade 2 | 14 | 15 | 19 | 1.000 | 0.396 | 0.567 |

**B.4 GLM 10-shot ICL**

| True \ Pred | grade 0 | grade 1 | grade 2 | Precision | Recall | F1 |
|---|---|---|---|---|---|---|
| grade 0 | 189 | 27 | 4 | 0.784 | 0.859 | 0.820 |
| grade 1 | 22 | 26 | 2 | 0.248 | 0.520 | 0.335 |
| grade 2 | 30 | 52 | 158 | 0.963 | 0.658 | 0.782 |

**B.5 Qwen 10-shot ICL**

| True \ Pred | grade 0 | grade 1 | grade 2 | Precision | Recall | F1 |
|---|---|---|---|---|---|---|
| grade 0 | 195 | 19 | 6 | 0.786 | 0.886 | 0.833 |
| grade 1 | 21 | 25 | 4 | 0.294 | 0.500 | 0.370 |
| grade 2 | 32 | 41 | 167 | 0.944 | 0.696 | 0.801 |

**B.6 Gemma 10-shot ICL**

| True \ Pred | grade 0 | grade 1 | grade 2 | Precision | Recall | F1 |
|---|---|---|---|---|---|---|
| grade 0 | 202 | 17 | 1 | 0.716 | 0.918 | 0.805 |
| grade 1 | 36 | 14 | 0 | 0.151 | 0.280 | 0.196 |
| grade 2 | 44 | 62 | 134 | 0.993 | 0.558 | 0.715 |

**B.7 CLIP linear-probe, 10 shot**

| True \ Pred | grade 0 | grade 1 | grade 2 | Precision | Recall | F1 |
|---|---|---|---|---|---|---|
| grade 0 | 128 | 60 | 32 | 0.800 | 0.582 | 0.674 |
| grade 1 | 11 | 24 | 15 | 0.170 | 0.480 | 0.251 |
| grade 2 | 21 | 57 | 162 | 0.775 | 0.675 | 0.722 |

**B.8 SigLIP2 linear-probe, 10 shot**

| True \ Pred | grade 0 | grade 1 | grade 2 | Precision | Recall | F1 |
|---|---|---|---|---|---|---|
| grade 0 | 161 | 37 | 22 | 0.605 | 0.732 | 0.663 |
| grade 1 | 28 | 15 | 7 | 0.128 | 0.300 | 0.180 |
| grade 2 | 77 | 65 | 98 | 0.772 | 0.408 | 0.534 |

**B.9 EyeCLIP linear-probe, 10 shot**

| True \ Pred | grade 0 | grade 1 | grade 2 | Precision | Recall | F1 |
|---|---|---|---|---|---|---|
| grade 0 | 42 | 84 | 94 | 0.442 | 0.191 | 0.267 |
| grade 1 | 8 | 18 | 24 | 0.093 | 0.360 | 0.148 |
| grade 2 | 45 | 92 | 103 | 0.466 | 0.429 | 0.447 |

**B.10 DINOv3 linear-probe, 10 shot**

| True \ Pred | grade 0 | grade 1 | grade 2 | Precision | Recall | F1 |
|---|---|---|---|---|---|---|
| grade 0 | 99 | 75 | 46 | 0.756 | 0.450 | 0.564 |
| grade 1 | 6 | 21 | 23 | 0.136 | 0.420 | 0.206 |
| grade 2 | 26 | 58 | 156 | 0.693 | 0.650 | 0.671 |

**B.11 VisionFM linear-probe, 10 shot**

| True \ Pred | grade 0 | grade 1 | grade 2 | Precision | Recall | F1 |
|---|---|---|---|---|---|---|
| grade 0 | 127 | 77 | 16 | 0.722 | 0.577 | 0.641 |
| grade 1 | 9 | 28 | 13 | 0.164 | 0.560 | 0.253 |
| grade 2 | 40 | 66 | 134 | 0.822 | 0.558 | 0.665 |

**B.12 RETFound linear-probe, 10 shot**

| True \ Pred | grade 0 | grade 1 | grade 2 | Precision | Recall | F1 |
|---|---|---|---|---|---|---|
| grade 0 | 92 | 42 | 86 | 0.676 | 0.418 | 0.517 |
| grade 1 | 12 | 15 | 23 | 0.118 | 0.300 | 0.169 |
| grade 2 | 32 | 70 | 138 | 0.559 | 0.575 | 0.567 |

**B.13 Bootstrapped metrics, IDRiD DME (all bootstrapped metrics)**

| Arm | accuracy | macro f1 | qwk | balanced accuracy |
|---|---|---|---|---|
| GLM zero-shot | 0.794 (0.716-0.873) | 0.641 (0.544-0.745) | 0.776 (0.660-0.874) | 0.642 (0.555-0.749) |
| Qwen zero-shot | 0.657 (0.569-0.745) | 0.529 (0.442-0.617) | 0.605 (0.469-0.731) | 0.545 (0.452-0.651) |
| Gemma zero-shot | 0.618 (0.520-0.716) | 0.494 (0.407-0.585) | 0.546 (0.412-0.679) | 0.517 (0.427-0.624) |
| CLIP zero-shot | 0.098 (0.049-0.157) | 0.060 (0.031-0.090) | 0.000 (0.000-0.000) | 0.333 (0.333-0.333) |
| SigLIP2 zero-shot | 0.471 (0.373-0.569) | 0.213 (0.181-0.242) | 0.000 (0.000-0.000) | 0.333 (0.333-0.333) |
| EyeCLIP zero-shot | 0.412 (0.314- | 0.236 (0.170- | -0.003 (-0.052-0.046) | 0.344 (0.291- |

| | | | | |
|---|---|---|---|---|
| | 0.510) | 0.322) | | 0.426) |
| GLM 5-shot ICL | 0.761 (0.684-0.831) | 0.657 (0.565-0.739) | 0.735 (0.614-0.838) | 0.676 (0.577-0.775) |
| Qwen 5-shot ICL | 0.733 (0.649-0.812) | 0.634 (0.543-0.718) | 0.709 (0.583-0.821) | 0.655 (0.553-0.760) |
| Gemma 5-shot ICL | 0.647 (0.559-0.731) | 0.533 (0.452-0.614) | 0.610 (0.484-0.724) | 0.547 (0.455-0.654) |
| CLIP linear-probe, 5 shot | 0.582 (0.518-0.645) | 0.513 (0.455-0.565) | 0.530 (0.409-0.635) | 0.528 (0.470-0.586) |
| CLIP partial-FT (last-2 layers), 5 shot | 0.598 (0.531-0.665) | 0.541 (0.474-0.601) | 0.556 (0.431-0.666) | 0.577 (0.505-0.648) |
| SigLIP2 linear-probe, 5 shot | 0.449 (0.398-0.500) | 0.397 (0.349-0.441) | 0.293 (0.203-0.378) | 0.442 (0.397-0.485) |
| SigLIP2 partial-FT (last-2 layers), 5 shot | 0.471 (0.410-0.531) | 0.447 (0.386-0.505) | 0.417 (0.307-0.516) | 0.513 (0.447-0.574) |
| EyeCLIP linear-probe, 5 shot | 0.294 (0.271-0.318) | 0.275 (0.247-0.299) | -0.041 (-0.091-0.010) | 0.306 (0.279-0.331) |
| EyeCLIP partial-FT (last-2 layers), 5 shot | 0.322 (0.300-0.343) | 0.293 (0.268-0.315) | 0.002 (-0.042-0.046) | 0.311 (0.286-0.339) |
| DINOv3 linear-probe, 5 shot | 0.449 (0.398-0.498) | 0.427 (0.373-0.476) | 0.369 (0.259-0.465) | 0.477 (0.414-0.535) |
| DINOv3 partial-FT (last-2 layers), 5 shot | 0.435 (0.384-0.484) | 0.418 (0.365-0.466) | 0.367 (0.259-0.462) | 0.472 (0.422-0.522) |
| VisionFM linear-probe, 5 shot | 0.531 (0.476-0.586) | 0.486 (0.432-0.534) | 0.475 (0.373-0.569) | 0.518 (0.462-0.573) |
| VisionFM partial-FT (last-2 layers), 5 shot | 0.561 (0.506-0.616) | 0.501 (0.450-0.546) | 0.494 (0.392-0.586) | 0.524 (0.473-0.578) |
| RETFound linear-probe, 5 shot | 0.325 (0.280-0.369) | 0.313 (0.265-0.359) | 0.143 (0.076-0.207) | 0.379 (0.305-0.446) |
| RETFound partial-FT (last-2 layers), 5 shot | 0.329 (0.286-0.375) | 0.320 (0.273-0.366) | 0.156 (0.084-0.223) | 0.383 (0.308-0.449) |
| GLM 10-shot ICL | 0.731 (0.653-0.804) | 0.646 (0.563-0.722) | 0.726 (0.608-0.828) | 0.679 (0.584-0.773) |
| Qwen 10-shot ICL | 0.759 (0.680-0.833) | 0.668 (0.578-0.751) | 0.734 (0.611-0.841) | 0.694 (0.595-0.789) |
| Gemma 10-shot ICL | 0.686 (0.602-0.769) | 0.572 (0.492-0.655) | 0.668 (0.543-0.783) | 0.586 (0.492-0.690) |
| CLIP linear-probe, 10 shot | 0.616 (0.541-0.688) | 0.549 (0.478-0.614) | 0.570 (0.437-0.683) | 0.579 (0.504-0.652) |
| CLIP partial-FT (last-2 layers), 10 shot | 0.575 (0.498-0.651) | 0.542 (0.469-0.611) | 0.619 (0.507-0.715) | 0.602 (0.525-0.676) |
| SigLIP2 linear-probe, 10 shot | 0.537 (0.471-0.604) | 0.459 (0.398-0.515) | 0.383 (0.257-0.502) | 0.480 (0.411-0.549) |
| SigLIP2 partial-FT (last-2 layers), 10 shot | 0.582 (0.514-0.649) | 0.520 (0.456-0.580) | 0.546 (0.432-0.641) | 0.553 (0.474-0.630) |
| EyeCLIP linear-probe, 10 shot | 0.320 (0.292-0.347) | 0.287 (0.261-0.309) | 0.003 (-0.037-0.043) | 0.327 (0.304-0.346) |
| EyeCLIP partial-FT (last-2 layers), 10 shot | 0.361 (0.331-0.388) | 0.309 (0.287-0.329) | 0.010 (-0.026-0.046) | 0.326 (0.306-0.348) |
| DINOv3 linear-probe, 10 shot | 0.541 (0.476-0.606) | 0.480 (0.419-0.538) | 0.444 (0.319-0.552) | 0.507 (0.434-0.584) |
| DINOv3 partial-FT (last-2 layers), 10 shot | 0.533 (0.469-0.596) | 0.485 (0.423-0.543) | 0.470 (0.359-0.566) | 0.527 (0.449-0.604) |
| VisionFM linear-probe, 10 shot | 0.567 (0.502- | 0.520 (0.456- | 0.514 (0.397-0.616) | 0.565 (0.501- |

| | | | | |
|---|---|---|---|---|
| | 0.631) | 0.579) | | 0.629) |
| VisionFM partial-FT (last-2 layers), 10 shot | 0.584 (0.518-0.651) | 0.528 (0.464-0.588) | 0.543 (0.430-0.645) | 0.562 (0.487-0.635) |
| RETFound linear-probe, 10 shot | 0.480 (0.425-0.535) | 0.418 (0.367-0.465) | 0.258 (0.163-0.347) | 0.431 (0.371-0.488) |
| RETFound partial-FT (last-2 layers), 10 shot | 0.484 (0.427-0.539) | 0.439 (0.379-0.494) | 0.297 (0.202-0.386) | 0.471 (0.395-0.547) |
| CLIP linear-probe, full dataset | 0.775 (0.696-0.853) | 0.625 (0.532-0.729) | 0.745 (0.617-0.856) | 0.624 (0.534-0.734) |
| CLIP partial-FT (last-2 layers), full dataset | 0.765 (0.676-0.843) | 0.656 (0.550-0.757) | 0.736 (0.611-0.846) | 0.674 (0.560-0.794) |
| SigLIP2 linear-probe, full dataset | 0.725 (0.637-0.814) | 0.514 (0.459-0.564) | 0.604 (0.448-0.743) | 0.537 (0.481-0.588) |
| SigLIP2 partial-FT (last-2 layers), full dataset | 0.824 (0.745-0.892) | 0.578 (0.534-0.616) | 0.785 (0.667-0.887) | 0.610 (0.571-0.645) |
| EyeCLIP linear-probe, full dataset | 0.529 (0.431-0.627) | 0.358 (0.290-0.422) | 0.151 (-0.032-0.330) | 0.386 (0.322-0.448) |
| EyeCLIP partial-FT (last-2 layers), full dataset | 0.549 (0.451-0.647) | 0.371 (0.300-0.434) | 0.191 (0.010-0.362) | 0.400 (0.337-0.459) |
| DINOv3 linear-probe, full dataset | 0.784 (0.706-0.863) | 0.637 (0.529-0.748) | 0.724 (0.588-0.842) | 0.634 (0.544-0.741) |
| DINOv3 partial-FT (last-2 layers), full dataset | 0.784 (0.706-0.863) | 0.558 (0.509-0.600) | 0.745 (0.617-0.854) | 0.582 (0.536-0.623) |
| VisionFM linear-probe, full dataset | 0.735 (0.647-0.824) | 0.569 (0.472-0.687) | 0.594 (0.436-0.738) | 0.571 (0.497-0.661) |
| VisionFM partial-FT (last-2 layers), full dataset | 0.775 (0.686-0.853) | 0.633 (0.518-0.747) | 0.687 (0.537-0.813) | 0.625 (0.533-0.732) |
| RETFound linear-probe, full dataset | 0.755 (0.667-0.833) | 0.535 (0.481-0.581) | 0.667 (0.519-0.794) | 0.559 (0.507-0.607) |
| RETFound partial-FT (last-2 layers), full dataset | 0.765 (0.676-0.843) | 0.536 (0.483-0.584) | 0.659 (0.509-0.794) | 0.566 (0.515-0.612) |

**Figure 2, Panel C - CFP Cup/Disc Localization (REFUGE)**

Dataset size: train+val n = 400 images; test n = 400 images.

**C.1 Bootstrapped metrics, REFUGE vCDR MAE**

| Arm | vcdr mae |
|---|---|
| GLM zero-shot | 0.104 (0.096-0.111) |
| Qwen zero-shot | 0.106 (0.099-0.113) |
| Gemma zero-shot | 0.150 (0.141-0.158) |
| GLM 5-shot ICL | 0.163 (0.155-0.171) |
| Qwen 5-shot ICL | 0.081 (0.076-0.086) |
| Gemma 5-shot ICL | 0.147 (0.140-0.154) |
| SigLIP2 linear-probe, 5 shot | 0.218 (0.210-0.228) |
| DINOv3 linear-probe, 5 shot | 0.374 (0.364-0.384) |
| VisionFM linear-probe, 5 shot | 0.252 (0.244-0.260) |
| RETFound linear-probe, 5 shot | 0.223 (0.217-0.228) |
| GLM 10-shot ICL | 0.169 (0.162-0.177) |
| Qwen 10-shot ICL | 0.087 (0.082-0.092) |
| Gemma 10-shot ICL | 0.122 (0.116-0.128) |
| CLIP linear-probe, 10 shot | 0.304 (0.288-0.323) |

| | |
|---|---|
| SigLIP2 linear-probe, 10 shot | 0.195 (0.189-0.201) |
| DINOv3 linear-probe, 10 shot | 0.421 (0.411-0.431) |
| VisionFM linear-probe, 10 shot | 0.201 (0.195-0.207) |
| RETFound linear-probe, 10 shot | 0.140 (0.135-0.145) |
| CLIP linear-probe, full dataset | 0.083 (0.077-0.089) |
| CLIP partial-FT (last-2 layers), full dataset | 0.064 (0.059-0.069) |
| CLIP full-FT, full dataset | 0.050 (0.047-0.054) |
| SigLIP2 linear-probe, full dataset | 0.082 (0.076-0.088) |
| SigLIP2 partial-FT (last-2 layers), full dataset | 0.070 (0.065-0.075) |
| SigLIP2 full-FT, full dataset | 0.065 (0.061-0.070) |
| EyeCLIP linear-probe, full dataset | 0.303 (0.264-0.343) |
| EyeCLIP partial-FT (last-2 layers), full dataset | 0.270 (0.247-0.294) |
| EyeCLIP full-FT, full dataset | 0.144 (0.134-0.153) |
| DINOv3 linear-probe, full dataset | 0.074 (0.068-0.079) |
| DINOv3 partial-FT (last-2 layers), full dataset | 0.071 (0.067-0.076) |
| DINOv3 full-FT, full dataset | 0.075 (0.070-0.080) |
| VisionFM linear-probe, full dataset | 0.115 (0.107-0.123) |
| VisionFM partial-FT (last-2 layers), full dataset | 0.114 (0.107-0.120) |
| VisionFM full-FT, full dataset | 0.082 (0.077-0.087) |
| RETFound linear-probe, full dataset | 0.092 (0.087-0.097) |
| RETFound partial-FT (last-2 layers), full dataset | 0.081 (0.076-0.086) |
| RETFound full-FT, full dataset | 0.078 (0.073-0.082) |

### C.2 Bootstrapped metrics, REFUGE glaucoma screening AUROC

| Arm | screening auroc |
|---|---|
| GLM zero-shot | 0.810 (0.730-0.882) |
| Qwen zero-shot | 0.828 (0.751-0.894) |
| Gemma zero-shot | 0.820 (0.745-0.888) |
| zs-clip zero-shot | 0.423 (0.340-0.507) |
| zs-eyeclip zero-shot | 0.488 (0.390-0.588) |
| zs-siglip2 zero-shot | 0.403 (0.300-0.504) |
| zs-clip descriptive 0 | 0.604 (0.508-0.696) |
| zs-clip zero_shot_ensemble 0 | 0.548 (0.443-0.651) |
| zs-eyeclip descriptive 0 | 0.482 (0.382-0.585) |
| zs-eyeclip zero_shot_ensemble 0 | 0.581 (0.468-0.692) |
| zs-siglip2 descriptive 0 | 0.467 (0.370-0.566) |
| zs-siglip2 zero_shot_ensemble 0 | 0.470 (0.375-0.564) |
| GLM 5-shot ICL | 0.783 (0.709-0.848) |
| Qwen 5-shot ICL | 0.822 (0.755-0.882) |
| Gemma 5-shot ICL | 0.820 (0.758-0.878) |
| CLIP linear-probe, 5 shot | 0.636 (0.599-0.671) |
| SigLIP2 linear-probe, 5 shot | 0.563 (0.518-0.609) |
| EyeCLIP linear-probe, 5 shot | 0.511 (0.467-0.554) |
| DINOv3 linear-probe, 5 shot | 0.544 (0.516-0.570) |
| VisionFM linear-probe, 5 shot | 0.833 (0.786-0.875) |
| RETFound linear-probe, 5 shot | 0.628 (0.585-0.668) |
| GLM 10-shot ICL | 0.785 (0.712-0.851) |

| | |
|---|---|
| Qwen 10-shot ICL | 0.802 (0.732-0.868) |
| Gemma 10-shot ICL | 0.816 (0.752-0.872) |
| CLIP linear-probe, 10 shot | 0.679 (0.630-0.721) |
| SigLIP2 linear-probe, 10 shot | 0.578 (0.540-0.619) |
| EyeCLIP linear-probe, 10 shot | 0.379 (0.277-0.465) |
| DINOv3 linear-probe, 10 shot | 0.602 (0.551-0.653) |
| VisionFM linear-probe, 10 shot | 0.830 (0.783-0.872) |
| RETFound linear-probe, 10 shot | 0.629 (0.571-0.682) |
| CLIP linear-probe, full dataset | 0.801 (0.698-0.892) |
| CLIP partial-FT (last-2 layers), full dataset | 0.849 (0.762-0.924) |
| CLIP full-FT, full dataset | 0.951 (0.918-0.978) |
| SigLIP2 linear-probe, full dataset | 0.848 (0.781-0.906) |
| SigLIP2 partial-FT (last-2 layers), full dataset | 0.906 (0.851-0.952) |
| SigLIP2 full-FT, full dataset | 0.937 (0.896-0.971) |
| EyeCLIP linear-probe, full dataset | 0.409 (0.169-0.672) |
| EyeCLIP partial-FT (last-2 layers), full dataset | 0.438 (0.322-0.557) |
| EyeCLIP full-FT, full dataset | 0.697 (0.602-0.788) |
| DINOv3 linear-probe, full dataset | 0.841 (0.761-0.914) |
| DINOv3 partial-FT (last-2 layers), full dataset | 0.892 (0.829-0.944) |
| DINOv3 full-FT, full dataset | 0.899 (0.826-0.958) |
| VisionFM linear-probe, full dataset | 0.922 (0.863-0.969) |
| VisionFM partial-FT (last-2 layers), full dataset | 0.934 (0.887-0.973) |
| VisionFM full-FT, full dataset | 0.956 (0.923-0.982) |
| RETFound linear-probe, full dataset | 0.935 (0.878-0.974) |
| RETFound partial-FT (last-2 layers), full dataset | 0.970 (0.936-0.993) |
| RETFound full-FT, full dataset | 0.947 (0.897-0.982) |

AUROC is threshold-independent: the predicted vCDR ranks against the binary glaucoma label (Mann-Whitney U); no threshold is applied. 95% CIs use the same image-level bootstrap.

**Figure 2, Panel D - Cross-dataset Cup/Disc Localization (REFUGE→OHTS)**

Dataset size: no training data (external validation); test n = 1,390 images.

**D.1 Bootstrapped metrics, OHTS vCDR MAE**

| Arm | vcdr mae |
|---|---|
| GLM zero-shot | 0.141 (0.135-0.147) |
| Qwen zero-shot | 0.152 (0.145-0.160) |
| Gemma zero-shot | 0.160 (0.153-0.168) |
| CLIP linear-probe, 5 shot | 2.719 (2.371-3.096) |
| SigLIP2 linear-probe, 5 shot | 0.510 (0.449-0.583) |
| EyeCLIP linear-probe, 5 shot | 1.949 (1.662-2.307) |
| DINOv3 linear-probe, 5 shot | 0.448 (0.411-0.494) |
| VisionFM linear-probe, 5 shot | 0.260 (0.253-0.267) |
| RETFound linear-probe, 5 shot | 0.370 (0.353-0.389) |
| GLM 10-shot ICL | 0.150 (0.144-0.157) |
| Qwen 10-shot ICL | 0.164 (0.157-0.171) |
| Gemma 10-shot ICL | 0.153 (0.147-0.161) |

| | |
|---|---|
| CLIP linear-probe, 10 shot | 3.994 (3.549-4.475) |
| SigLIP2 linear-probe, 10 shot | 0.382 (0.342-0.431) |
| EyeCLIP linear-probe, 10 shot | 2.618 (1.983-3.373) |
| DINOv3 linear-probe, 10 shot | 0.529 (0.486-0.581) |
| VisionFM linear-probe, 10 shot | 0.208 (0.202-0.214) |
| RETFound linear-probe, 10 shot | 0.320 (0.310-0.329) |
| CLIP linear-probe, full dataset | 0.224 (0.209-0.240) |
| CLIP full-FT, full dataset | 0.178 (0.169-0.187) |
| CLIP partial-FT (last-2 layers), full dataset | 0.194 (0.185-0.204) |
| SigLIP2 linear-probe, full dataset | 0.294 (0.252-0.356) |
| SigLIP2 full-FT, full dataset | 0.278 (0.266-0.290) |
| SigLIP2 partial-FT (last-2 layers), full dataset | 0.216 (0.206-0.226) |
| EyeCLIP linear-probe, full dataset | 0.369 (0.339-0.407) |
| EyeCLIP full-FT, full dataset | 0.185 (0.175-0.196) |
| EyeCLIP partial-FT (last-2 layers), full dataset | 0.295 (0.282-0.309) |
| DINOv3 linear-probe, full dataset | 0.223 (0.212-0.234) |
| DINOv3 full-FT, full dataset | 0.224 (0.214-0.235) |
| DINOv3 partial-FT (last-2 layers), full dataset | 0.248 (0.236-0.261) |
| VisionFM linear-probe, full dataset | 0.184 (0.176-0.192) |
| VisionFM full-FT, full dataset | 0.152 (0.145-0.158) |
| VisionFM partial-FT (last-2 layers), full dataset | 0.197 (0.188-0.206) |
| RETFound linear-probe, full dataset | 0.305 (0.295-0.315) |
| RETFound full-FT, full dataset | 0.359 (0.346-0.373) |
| RETFound partial-FT (last-2 layers), full dataset | 0.380 (0.368-0.392) |

**D.2 OHTS vCDR MAE by demographic subgroup (patients n = 541, images n = 1,390)**

Subgroups were linked to baseline demographics using the frozen image manifest. Cell values represent per-image mean absolute error (MAE) pooled across random seeds, with 95% confidence intervals derived from paired, image-level bootstrapping (10,000 percentile replicates, seed 2026; replicate draws shared across arms within each subgroup). The "All" row reproduces the D.1 cohort.

Race classifications were defined using self-reported OHTS categories: "Black" and "White" denote non-Hispanic cohorts, while "Other" aggregates Hispanic, Asian/Pacific Islander, American Indian/Alaskan Native, and other designations. VisionFM was evaluated under two configurations transferred directly to OHTS without modification: a linear probe trained on 10 REFUGE shots and a model fine-tuned on the complete REFUGE dataset.

| Arm | Patients (n) | GLM 0-shot MAE | GLM 10-shot MAE | Qwen 0-shot MAE | Qwen 10-shot MAE | VisionFM 10-shot MAE | VisionFM fully-trained MAE |
|---|---|---|---|---|---|---|---|
| Overall: All | 541 | 0.141 (0.135-0.148) | 0.150 (0.144-0.157) | 0.152 (0.145-0.160) | 0.164 (0.157-0.171) | 0.208 (0.203-0.214) | 0.152 (0.145-0.158) |
| Race: Black | 140 | 0.127 (0.116-0.139) | 0.134 (0.122-0.145) | 0.136 (0.123-0.149) | 0.149 (0.136-0.162) | 0.189 (0.178-0.201) | 0.120 (0.110-0.131) |
| Race: White | 379 | 0.148 (0.141-0.156) | 0.158 (0.150-0.166) | 0.160 (0.152-0.169) | 0.171 (0.163-0.180) | 0.217 (0.210-0.224) | 0.166 (0.158-0.175) |
| Race: Other | 22 | 0.109 (0.087-0.133) | 0.116 (0.092-0.144) | 0.124 (0.096-0.156) | 0.140 (0.111-0.173) | 0.187 (0.160-0.217) | 0.098 (0.076-0.122) |

| | | | | | | | |
|---|---|---|---|---|---|---|---|
| Sex: Female | 323 | 0.147 (0.139-0.155) | 0.159 (0.150-0.168) | 0.158 (0.149-0.168) | 0.170 (0.161-0.180) | 0.214 (0.206-0.221) | 0.153 (0.145-0.162) |
| Sex: Male | 218 | 0.133 (0.124-0.142) | 0.136 (0.127-0.146) | 0.144 (0.133-0.155) | 0.155 (0.144-0.166) | 0.200 (0.191-0.210) | 0.149 (0.139-0.160) |
| Age at randomization: <60 years | 445 | 0.140 (0.133-0.147) | 0.150 (0.143-0.157) | 0.153 (0.145-0.161) | 0.164 (0.156-0.172) | 0.210 (0.204-0.217) | 0.152 (0.145-0.159) |
| Age at randomization: >=60 years | 96 | 0.147 (0.132-0.164) | 0.151 (0.135-0.167) | 0.150 (0.132-0.167) | 0.164 (0.146-0.183) | 0.201 (0.186-0.216) | 0.150 (0.134-0.167) |

**Figure 2, Panel E - OCT Classification (UCSD)**

Dataset size: train+val n = 108,309 images; test n = 1,000 images.

**E.1 GLM zero-shot**

| True \ Pred | CNV | DME | DRUSEN | NORMAL | Precision | Recall | F1 |
|---|---|---|---|---|---|---|---|
| CNV | 156 | 47 | 47 | 0 | 0.994 | 0.624 | 0.767 |
| DME | 1 | 247 | 0 | 2 | 0.840 | 0.988 | 0.908 |
| DRUSEN | 0 | 0 | 212 | 38 | 0.819 | 0.848 | 0.833 |
| NORMAL | 0 | 0 | 0 | 250 | 0.862 | 1.000 | 0.926 |

**E.2 Qwen zero-shot**

| True \ Pred | CNV | DME | DRUSEN | NORMAL | Precision | Recall | F1 |
|---|---|---|---|---|---|---|---|
| CNV | 139 | 54 | 57 | 0 | 1.000 | 0.556 | 0.715 |
| DME | 0 | 244 | 3 | 3 | 0.819 | 0.976 | 0.891 |
| DRUSEN | 0 | 0 | 226 | 24 | 0.771 | 0.904 | 0.832 |
| NORMAL | 0 | 0 | 7 | 243 | 0.900 | 0.972 | 0.935 |

**E.3 Gemma zero-shot**

| True \ Pred | CNV | DME | DRUSEN | NORMAL | Precision | Recall | F1 |
|---|---|---|---|---|---|---|---|
| CNV | 188 | 18 | 44 | 0 | 0.940 | 0.752 | 0.836 |
| DME | 12 | 228 | 5 | 5 | 0.927 | 0.912 | 0.919 |
| DRUSEN | 0 | 0 | 225 | 25 | 0.784 | 0.900 | 0.838 |
| NORMAL | 0 | 0 | 13 | 237 | 0.888 | 0.948 | 0.917 |

**E.4 GLM 10-shot ICL**

| True \ Pred | CNV | DME | DRUSEN | NORMAL | Precision | Recall | F1 |
|---|---|---|---|---|---|---|---|
| CNV | 946 | 136 | 167 | 1 | 0.976 | 0.757 | 0.853 |
| DME | 15 | 1235 | 0 | 0 | 0.899 | 0.988 | 0.942 |
| DRUSEN | 8 | 1 | 1087 | 154 | 0.866 | 0.870 | 0.868 |
| NORMAL | 0 | 1 | 1 | 1248 | 0.890 | 0.998 | 0.941 |

**E.5 Qwen 10-shot ICL**

| True \ Pred | CNV | DME | DRUSEN | NORMAL | Precision | Recall | F1 |
|---|---|---|---|---|---|---|---|
| CNV | 1084 | 107 | 59 | 0 | 0.973 | 0.867 | 0.917 |
| DME | 11 | 1238 | 1 | 0 | 0.910 | 0.990 | 0.948 |
| DRUSEN | 19 | 6 | 1146 | 79 | 0.939 | 0.917 | 0.928 |
| NORMAL | 0 | 10 | 14 | 1226 | 0.939 | 0.981 | 0.960 |

**E.6 Gemma 10-shot ICL**

| True \ Pred | CNV | DME | DRUSEN | NORMAL | Precision | Recall | F1 |
|---|---|---|---|---|---|---|---|
| CNV | 1024 | 73 | 149 | 4 | 0.944 | 0.819 | 0.877 |
| DME | 55 | 1173 | 7 | 15 | 0.941 | 0.938 | 0.940 |
| DRUSEN | 6 | 0 | 975 | 269 | 0.860 | 0.780 | 0.818 |
| NORMAL | 0 | 0 | 3 | 1247 | 0.812 | 0.998 | 0.896 |

**E.7 CLIP linear-probe, 10 shot**

| True \ Pred | CNV | DME | DRUSEN | NORMAL | Precision | Recall | F1 |
|---|---|---|---|---|---|---|---|
| CNV | 479 | 592 | 92 | 87 | 0.326 | 0.383 | 0.352 |
| DME | 358 | 739 | 68 | 85 | 0.320 | 0.591 | 0.415 |
| DRUSEN | 410 | 474 | 172 | 194 | 0.390 | 0.138 | 0.203 |
| NORMAL | 224 | 505 | 109 | 412 | 0.530 | 0.330 | 0.406 |

**E.8 SigLIP2 linear-probe, 10 shot**

| True \ Pred | CNV | DME | DRUSEN | NORMAL | Precision | Recall | F1 |
|---|---|---|---|---|---|---|---|
| CNV | 452 | 431 | 117 | 250 | 0.439 | 0.362 | 0.396 |
| DME | 171 | 733 | 54 | 292 | 0.402 | 0.586 | 0.477 |
| DRUSEN | 330 | 286 | 269 | 365 | 0.434 | 0.215 | 0.288 |
| NORMAL | 77 | 372 | 180 | 621 | 0.406 | 0.497 | 0.447 |

**E.9 EyeCLIP linear-probe, 10 shot**

| True \ Pred | CNV | DME | DRUSEN | NORMAL | Precision | Recall | F1 |
|---|---|---|---|---|---|---|---|
| CNV | 152 | 71 | 401 | 626 | 0.304 | 0.122 | 0.174 |
| DME | 147 | 67 | 423 | 613 | 0.191 | 0.054 | 0.084 |
| DRUSEN | 125 | 96 | 436 | 593 | 0.249 | 0.349 | 0.291 |
| NORMAL | 76 | 117 | 490 | 567 | 0.236 | 0.454 | 0.311 |

**E.10 DINOv3 linear-probe, 10 shot**

| True \ Pred | CNV | DME | DRUSEN | NORMAL | Precision | Recall | F1 |
|---|---|---|---|---|---|---|---|
| CNV | 587 | 172 | 351 | 140 | 0.611 | 0.470 | 0.531 |
| DME | 123 | 694 | 115 | 318 | 0.691 | 0.555 | 0.616 |
| DRUSEN | 240 | 91 | 590 | 329 | 0.517 | 0.472 | 0.493 |
| NORMAL | 11 | 47 | 86 | 1106 | 0.584 | 0.885 | 0.704 |

**E.11 VisionFM linear-probe, 10 shot**

| True \ Pred | CNV | DME | DRUSEN | NORMAL | Precision | Recall | F1 |
|---|---|---|---|---|---|---|---|
| CNV | 542 | 30 | 264 | 414 | 0.353 | 0.434 | 0.389 |
| DME | 439 | 26 | 310 | 475 | 0.342 | 0.021 | 0.039 |
| DRUSEN | 329 | 14 | 391 | 516 | 0.300 | 0.313 | 0.306 |
| NORMAL | 225 | 6 | 340 | 679 | 0.326 | 0.543 | 0.407 |

**E.12 RETFound linear-probe, 10 shot**

| True \ Pred | CNV | DME | DRUSEN | NORMAL | Precision | Recall | F1 |
|---|---|---|---|---|---|---|---|
| CNV | 623 | 160 | 323 | 144 | 0.289 | 0.498 | 0.366 |
| DME | 590 | 155 | 354 | 151 | 0.334 | 0.124 | 0.181 |

| | | | | | | | |
|---|---|---|---|---|---|---|---|
| DRUSEN | 539 | 103 | 433 | 175 | 0.261 | 0.346 | 0.298 |
| NORMAL | 402 | 46 | 549 | 253 | 0.350 | 0.202 | 0.256 |

**E.13 CLIP linear-probe, full dataset**

| True \ Pred | CNV | DME | DRUSEN | NORMAL | Precision | Recall | F1 |
|---|---|---|---|---|---|---|---|
| CNV | 243 | 4 | 1 | 2 | 0.620 | 0.972 | 0.757 |
| DME | 46 | 202 | 0 | 2 | 0.957 | 0.808 | 0.876 |
| DRUSEN | 102 | 4 | 105 | 39 | 0.938 | 0.420 | 0.580 |
| NORMAL | 1 | 1 | 6 | 242 | 0.849 | 0.968 | 0.905 |

**E.14 SigLIP2 linear-probe, full dataset**

| True \ Pred | CNV | DME | DRUSEN | NORMAL | Precision | Recall | F1 |
|---|---|---|---|---|---|---|---|
| CNV | 243 | 4 | 3 | 0 | 0.799 | 0.972 | 0.877 |
| DME | 39 | 209 | 0 | 2 | 0.929 | 0.836 | 0.880 |
| DRUSEN | 21 | 9 | 193 | 27 | 0.946 | 0.772 | 0.850 |
| NORMAL | 1 | 3 | 8 | 238 | 0.891 | 0.952 | 0.921 |

**E.15 EyeCLIP linear-probe, full dataset**

| True \ Pred | CNV | DME | DRUSEN | NORMAL | Precision | Recall | F1 |
|---|---|---|---|---|---|---|---|
| CNV | 26 | 0 | 0 | 224 | 0.306 | 0.104 | 0.155 |
| DME | 25 | 0 | 0 | 225 | 0.000 | 0.000 | 0.000 |
| DRUSEN | 18 | 0 | 0 | 232 | 0.000 | 0.000 | 0.000 |
| NORMAL | 16 | 0 | 0 | 234 | 0.256 | 0.936 | 0.402 |

**E.16 DINOv3 linear-probe, full dataset**

| True \ Pred | CNV | DME | DRUSEN | NORMAL | Precision | Recall | F1 |
|---|---|---|---|---|---|---|---|
| CNV | 247 | 3 | 0 | 0 | 0.789 | 0.988 | 0.877 |
| DME | 21 | 229 | 0 | 0 | 0.974 | 0.916 | 0.944 |
| DRUSEN | 44 | 1 | 198 | 7 | 0.980 | 0.792 | 0.876 |
| NORMAL | 1 | 2 | 4 | 243 | 0.972 | 0.972 | 0.972 |

**E.17 VisionFM linear-probe, full dataset**

| True \ Pred | CNV | DME | DRUSEN | NORMAL | Precision | Recall | F1 |
|---|---|---|---|---|---|---|---|
| CNV | 247 | 1 | 1 | 1 | 0.592 | 0.988 | 0.741 |
| DME | 72 | 162 | 1 | 15 | 0.982 | 0.648 | 0.781 |
| DRUSEN | 95 | 1 | 135 | 19 | 0.951 | 0.540 | 0.689 |
| NORMAL | 3 | 1 | 5 | 241 | 0.873 | 0.964 | 0.916 |

**E.18 RETFound linear-probe, full dataset**

| True \ Pred | CNV | DME | DRUSEN | NORMAL | Precision | Recall | F1 |
|---|---|---|---|---|---|---|---|
| CNV | 235 | 5 | 5 | 5 | 0.550 | 0.940 | 0.694 |
| DME | 112 | 58 | 14 | 66 | 0.817 | 0.232 | 0.361 |
| DRUSEN | 75 | 5 | 77 | 93 | 0.733 | 0.308 | 0.434 |
| NORMAL | 5 | 3 | 9 | 233 | 0.587 | 0.932 | 0.720 |

**E.19 Bootstrapped metrics, UCSD OCT**

| Arm | macro f1 | accuracy |
|---|---|---|

| | | |
|---|---|---|
| GLM zero-shot | 0.858 (0.836-0.880) | 0.865 (0.844-0.886) |
| Qwen zero-shot | 0.843 (0.820-0.865) | 0.852 (0.830-0.874) |
| Gemma zero-shot | 0.877 (0.857-0.897) | 0.878 (0.858-0.898) |
| CLIP zero-shot | 0.156 (0.140-0.173) | 0.241 (0.215-0.268) |
| CLIP descriptive 0 | 0.116 (0.103-0.131) | 0.251 (0.225-0.279) |
| CLIP zero_shot_ensemble 0 | 0.148 (0.131-0.165) | 0.257 (0.230-0.284) |
| SigLIP2 zero-shot | 0.100 (0.092-0.109) | 0.250 (0.223-0.277) |
| SigLIP2 descriptive 0 | 0.102 (0.093-0.111) | 0.249 (0.223-0.276) |
| SigLIP2 zero_shot_ensemble 0 | 0.166 (0.150-0.182) | 0.250 (0.223-0.277) |
| EyeCLIP zero-shot | 0.100 (0.092-0.108) | 0.250 (0.224-0.277) |
| EyeCLIP descriptive 0 | 0.104 (0.094-0.115) | 0.252 (0.225-0.279) |
| EyeCLIP zero_shot_ensemble 0 | 0.100 (0.091-0.108) | 0.250 (0.223-0.277) |
| GLM 5-shot ICL | 0.903 (0.886-0.919) | 0.906 (0.889-0.922) |
| Qwen 5-shot ICL | 0.946 (0.934-0.957) | 0.947 (0.934-0.958) |
| Gemma 5-shot ICL | 0.895 (0.877-0.911) | 0.895 (0.878-0.913) |
| CLIP linear-probe, 5 shot | 0.324 (0.313-0.335) | 0.329 (0.317-0.341) |
| SigLIP2 linear-probe, 5 shot | 0.366 (0.353-0.379) | 0.378 (0.364-0.393) |
| EyeCLIP linear-probe, 5 shot | 0.207 (0.197-0.216) | 0.240 (0.228-0.253) |
| DINOv3 linear-probe, 5 shot | 0.540 (0.522-0.556) | 0.552 (0.532-0.571) |
| VisionFM linear-probe, 5 shot | 0.268 (0.259-0.277) | 0.297 (0.284-0.310) |
| RETFound linear-probe, 5 shot | 0.263 (0.253-0.273) | 0.266 (0.256-0.277) |
| GLM 10-shot ICL | 0.901 (0.884-0.916) | 0.903 (0.887-0.919) |
| Qwen 10-shot ICL | 0.938 (0.925-0.950) | 0.939 (0.926-0.951) |
| Gemma 10-shot ICL | 0.883 (0.865-0.899) | 0.884 (0.866-0.901) |
| CLIP linear-probe, 10 shot | 0.344 (0.330-0.358) | 0.360 (0.344-0.376) |
| SigLIP2 linear-probe, 10 shot | 0.402 (0.388-0.416) | 0.415 (0.399-0.431) |
| EyeCLIP linear-probe, 10 shot | 0.215 (0.206-0.223) | 0.244 (0.232-0.256) |
| DINOv3 linear-probe, 10 shot | 0.586 (0.568-0.604) | 0.595 (0.576-0.615) |
| VisionFM linear-probe, 10 shot | 0.285 (0.276-0.295) | 0.328 (0.313-0.342) |
| RETFound linear-probe, 10 shot | 0.275 (0.262-0.288) | 0.293 (0.278-0.308) |
| CLIP linear-probe, full dataset | 0.780 (0.754-0.804) | 0.792 (0.766-0.817) |
| CLIP partial-FT (last-2 layers), full dataset | 0.843 (0.821-0.864) | 0.850 (0.828-0.872) |
| CLIP full-FT, full dataset | 0.854 (0.833-0.874) | 0.860 (0.838-0.881) |
| SigLIP2 linear-probe, full dataset | 0.882 (0.861-0.902) | 0.883 (0.863-0.903) |
| SigLIP2 partial-FT (last-2 layers), full dataset | 0.937 (0.922-0.951) | 0.937 (0.921-0.951) |
| SigLIP2 full-FT, full dataset | 0.875 (0.854-0.895) | 0.880 (0.859-0.900) |
| EyeCLIP linear-probe, full dataset | 0.139 (0.123-0.156) | 0.260 (0.233-0.287) |
| EyeCLIP partial-FT (last-2 layers), full dataset | 0.693 (0.663-0.722) | 0.711 (0.682-0.740) |
| EyeCLIP full-FT, full dataset | 0.879 (0.859-0.898) | 0.879 (0.858-0.899) |
| DINOv3 linear-probe, full dataset | 0.917 (0.900-0.934) | 0.917 (0.899-0.934) |
| DINOv3 partial-FT (last-2 layers), full dataset | 0.907 (0.889-0.924) | 0.908 (0.889-0.925) |
| DINOv3 full-FT, full dataset | 0.951 (0.937-0.963) | 0.951 (0.937-0.964) |
| VisionFM linear-probe, full dataset | 0.782 (0.755-0.806) | 0.785 (0.759-0.810) |
| VisionFM partial-FT (last-2 layers), full dataset | 0.917 (0.900-0.934) | 0.916 (0.899-0.933) |
| VisionFM full-FT, full dataset | 0.957 (0.944-0.969) | 0.957 (0.944-0.969) |
| RETFound linear-probe, full dataset | 0.552 (0.520-0.583) | 0.603 (0.572-0.633) |

| | | |
|---|---|---|
| RETFound partial-FT (last-2 layers), full dataset | 0.890 (0.871-0.909) | 0.889 (0.869-0.908) |
| RETFound full-FT, full dataset | 0.953 (0.940-0.966) | 0.953 (0.940-0.966) |

**Figure 2, Panel F - OCT Localization (BPIE-DLS dataset)**
Dataset size: train+val n = 9,617 images; test n = 3,971 images.

**F.1 GLM zero-shot**

| True \ Pred | no lesion | lesion | Precision | Recall | F1 |
|---|---|---|---|---|---|
| no lesion | 12593 | 4989 | 0.797 | 0.716 | 0.755 |
| lesion | 3204 | 10982 | 0.688 | 0.774 | 0.728 |

**F.2 Qwen zero-shot**

| True \ Pred | no lesion | lesion | Precision | Recall | F1 |
|---|---|---|---|---|---|
| no lesion | 12832 | 4695 | 0.773 | 0.732 | 0.752 |
| lesion | 3775 | 10330 | 0.688 | 0.732 | 0.709 |

**F.3 Gemma zero-shot**

| True \ Pred | no lesion | lesion | Precision | Recall | F1 |
|---|---|---|---|---|---|
| no lesion | 5905 | 11677 | 0.872 | 0.336 | 0.485 |
| lesion | 866 | 13320 | 0.533 | 0.939 | 0.680 |

**F.4 CLIP linear-probe, 10 shot**

| True \ Pred | no lesion | lesion | Precision | Recall | F1 |
|---|---|---|---|---|---|
| no lesion | 51220 | 36690 | 0.774 | 0.583 | 0.665 |
| lesion | 14971 | 55959 | 0.604 | 0.789 | 0.684 |

**F.5 SigLIP2 linear-probe, 10 shot**

| True \ Pred | no lesion | lesion | Precision | Recall | F1 |
|---|---|---|---|---|---|
| no lesion | 57405 | 30505 | 0.774 | 0.653 | 0.708 |
| lesion | 16741 | 54189 | 0.640 | 0.764 | 0.696 |

**F.6 EyeCLIP linear-probe, 10 shot**

| True \ Pred | no lesion | lesion | Precision | Recall | F1 |
|---|---|---|---|---|---|
| no lesion | 132 | 87778 | 0.742 | 0.002 | 0.003 |
| lesion | 46 | 70884 | 0.447 | 0.999 | 0.617 |

**F.7 DINOv3 linear-probe, 10 shot**

| True \ Pred | no lesion | lesion | Precision | Recall | F1 |
|---|---|---|---|---|---|
| no lesion | 62530 | 25380 | 0.722 | 0.711 | 0.717 |
| lesion | 24065 | 46865 | 0.649 | 0.661 | 0.655 |

**F.8 VisionFM linear-probe, 10 shot**

| True \ Pred | no lesion | lesion | Precision | Recall | F1 |
|---|---|---|---|---|---|
| no lesion | 60096 | 27814 | 0.725 | 0.684 | 0.704 |
| lesion | 22740 | 48190 | 0.634 | 0.679 | 0.656 |

**F.9 RETFound linear-probe, 10 shot**

| True \ Pred | no lesion | lesion | Precision | Recall | F1 |
|---|---|---|---|---|---|

| | | | | | |
|---|---|---|---|---|---|
| no lesion | 59662 | 28248 | 0.641 | 0.679 | 0.659 |
| lesion | 33461 | 37469 | 0.570 | 0.528 | 0.548 |

**F.10 CLIP linear-probe, full dataset**

| True \ Pred | no lesion | lesion | Precision | Recall | F1 |
|---|---|---|---|---|---|
| no lesion | 13837 | 3745 | 0.832 | 0.787 | 0.809 |
| lesion | 2792 | 11394 | 0.753 | 0.803 | 0.777 |

**F.11 SigLIP2 linear-probe, full dataset**

| True \ Pred | no lesion | lesion | Precision | Recall | F1 |
|---|---|---|---|---|---|
| no lesion | 13655 | 3927 | 0.851 | 0.777 | 0.812 |
| lesion | 2389 | 11797 | 0.750 | 0.832 | 0.789 |

**F.12 EyeCLIP linear-probe, full dataset**

| True \ Pred | no lesion | lesion | Precision | Recall | F1 |
|---|---|---|---|---|---|
| no lesion | 13115 | 4467 | 0.753 | 0.746 | 0.750 |
| lesion | 4294 | 9892 | 0.689 | 0.697 | 0.693 |

**F.13 DINOv3 linear-probe, full dataset**

| True \ Pred | no lesion | lesion | Precision | Recall | F1 |
|---|---|---|---|---|---|
| no lesion | 13946 | 3636 | 0.853 | 0.793 | 0.822 |
| lesion | 2411 | 11775 | 0.764 | 0.830 | 0.796 |

**F.14 VisionFM linear-probe, full dataset**

| True \ Pred | no lesion | lesion | Precision | Recall | F1 |
|---|---|---|---|---|---|
| no lesion | 14888 | 2694 | 0.850 | 0.847 | 0.848 |
| lesion | 2626 | 11560 | 0.811 | 0.815 | 0.813 |

**F.15 RETFound linear-probe, full dataset**

| True \ Pred | no lesion | lesion | Precision | Recall | F1 |
|---|---|---|---|---|---|
| no lesion | 14203 | 3379 | 0.849 | 0.808 | 0.828 |
| lesion | 2532 | 11654 | 0.775 | 0.822 | 0.798 |

**F.16 Bootstrapped metrics, BPEI-DLS pooled lesion F1**

| Arm | lesion f1 |
|---|---|
| GLM zero-shot | 0.728 (0.722-0.735) |
| Qwen zero-shot | 0.709 (0.702-0.716) |
| Gemma zero-shot | 0.680 (0.673-0.686) |
| GLM 5-shot ICL | 0.733 (0.726-0.739) |
| CLIP linear-probe, 5 shot | 0.663 (0.658-0.669) |
| CLIP partial-FT (last-2 layers), 5 shot | 0.660 (0.655-0.666) |
| SigLIP2 linear-probe, 5 shot | 0.661 (0.655-0.666) |
| SigLIP2 partial-FT (last-2 layers), 5 shot | 0.662 (0.656-0.668) |
| EyeCLIP linear-probe, 5 shot | 0.422 (0.419-0.425) |
| EyeCLIP partial-FT (last-2 layers), 5 shot | 0.276 (0.275-0.277) |
| DINOv3 linear-probe, 5 shot | 0.640 (0.635-0.646) |
| DINOv3 partial-FT (last-2 layers), 5 shot | 0.631 (0.626-0.637) |

| | |
|---|---|
| VisionFM linear-probe, 5 shot | 0.640 (0.634-0.645) |
| VisionFM partial-FT (last-2 layers), 5 shot | 0.640 (0.634-0.645) |
| RETFound linear-probe, 5 shot | 0.437 (0.434-0.440) |
| RETFound partial-FT (last-2 layers), 5 shot | 0.322 (0.320-0.325) |
| GLM 10-shot ICL | 0.728 (0.722-0.734) |
| Qwen 10-shot ICL | 0.750 (0.744-0.756) |
| Gemma 10-shot ICL | 0.747 (0.741-0.753) |
| CLIP linear-probe, 10 shot | 0.684 (0.678-0.690) |
| CLIP partial-FT (last-2 layers), 10 shot | 0.666 (0.660-0.671) |
| SigLIP2 linear-probe, 10 shot | 0.696 (0.690-0.702) |
| SigLIP2 partial-FT (last-2 layers), 10 shot | 0.688 (0.682-0.694) |
| EyeCLIP linear-probe, 10 shot | 0.617 (0.612-0.623) |
| EyeCLIP partial-FT (last-2 layers), 10 shot | 0.614 (0.609-0.620) |
| DINOv3 linear-probe, 10 shot | 0.655 (0.648-0.661) |
| DINOv3 partial-FT (last-2 layers), 10 shot | 0.663 (0.657-0.669) |
| VisionFM linear-probe, 10 shot | 0.656 (0.651-0.661) |
| VisionFM partial-FT (last-2 layers), 10 shot | 0.665 (0.660-0.670) |
| RETFound linear-probe, 10 shot | 0.548 (0.544-0.553) |
| RETFound partial-FT (last-2 layers), 10 shot | 0.632 (0.627-0.637) |
| CLIP linear-probe, full dataset | 0.777 (0.771-0.783) |
| CLIP partial-FT (last-2 layers), full dataset | 0.801 (0.796-0.807) |
| SigLIP2 linear-probe, full dataset | 0.789 (0.783-0.795) |
| SigLIP2 partial-FT (last-2 layers), full dataset | 0.818 (0.813-0.824) |
| EyeCLIP linear-probe, full dataset | 0.693 (0.686-0.700) |
| EyeCLIP partial-FT (last-2 layers), full dataset | 0.772 (0.766-0.778) |
| DINOv3 linear-probe, full dataset | 0.796 (0.790-0.802) |
| DINOv3 partial-FT (last-2 layers), full dataset | 0.856 (0.851-0.860) |
| VisionFM linear-probe, full dataset | 0.813 (0.807-0.819) |
| VisionFM partial-FT (last-2 layers), full dataset | 0.832 (0.827-0.838) |
| RETFound linear-probe, full dataset | 0.798 (0.792-0.803) |
| RETFound partial-FT (last-2 layers), full dataset | 0.816 (0.811-0.822) |

**Figure 2, Panel G - AS-OCT Classification (ASOCT-MSDC)**

Dataset size: train+val n = 956 images; test n = 150 images.

Metric keys: PACG = primary angle-closure glaucoma (binary axis); cataract = cataract (binary axis); cls4 = the 4-class map (Normal / Cataract / Glaucoma / Glaucoma+Cataract).

**G.1 GLM zero-shot**

| True \ Pred | Normal | Cataract | Glaucoma | Glaucoma_C | Precision | Recall | F1 |
|---|---|---|---|---|---|---|---|
| Normal | 32 | 0 | 12 | 0 | 0.711 | 0.727 | 0.719 |
| Cataract | 12 | 5 | 38 | 13 | 1.000 | 0.074 | 0.137 |
| Glaucoma | 1 | 0 | 14 | 4 | 0.177 | 0.737 | 0.286 |
| Glaucoma_Catarac | 0 | 0 | 15 | 4 | 0.190 | 0.211 | 0.200 |

**G.2 Qwen zero-shot**

| True \ Pred | Normal | Cataract | Glaucoma | Glaucoma_C | Precision | Recall | F1 |
|---|---|---|---|---|---|---|---|
| Normal | 43 | 0 | 1 | 0 | 0.350 | 0.977 | 0.515 |

| | | | | | | | |
|---|---|---|---|---|---|---|---|
| Cataract | 53 | 1 | 14 | 0 | 1.000 | 0.015 | 0.029 |
| Glaucoma | 17 | 0 | 2 | 0 | 0.077 | 0.105 | 0.089 |
| Glaucoma_Catarac | 10 | 0 | 9 | 0 | 0.000 | 0.000 | 0.000 |

### G.3 Gemma zero-shot

| True \ Pred | Normal | Cataract | Glaucoma | Glaucoma_C | Precision | Recall | F1 |
|---|---|---|---|---|---|---|---|
| Normal | 29 | 0 | 15 | 0 | 0.537 | 0.659 | 0.592 |
| Cataract | 25 | 1 | 42 | 0 | 1.000 | 0.015 | 0.029 |
| Glaucoma | 0 | 0 | 19 | 0 | 0.202 | 1.000 | 0.336 |
| Glaucoma_Catarac | 0 | 0 | 18 | 1 | 1.000 | 0.053 | 0.100 |

### G.4 GLM 10-shot ICL

| True \ Pred | Normal | Cataract | Glaucoma | Glaucoma_C | Precision | Recall | F1 |
|---|---|---|---|---|---|---|---|
| Normal | 219 | 1 | 0 | 0 | 0.582 | 0.995 | 0.735 |
| Cataract | 126 | 101 | 90 | 23 | 0.878 | 0.297 | 0.444 |
| Glaucoma | 18 | 4 | 62 | 11 | 0.297 | 0.653 | 0.408 |
| Glaucoma_Catarac | 13 | 9 | 57 | 16 | 0.320 | 0.168 | 0.221 |

### G.5 Qwen 10-shot ICL

| True \ Pred | Normal | Cataract | Glaucoma | Glaucoma_C | Precision | Recall | F1 |
|---|---|---|---|---|---|---|---|
| Normal | 216 | 4 | 0 | 0 | 0.871 | 0.982 | 0.923 |
| Cataract | 32 | 157 | 30 | 121 | 0.818 | 0.462 | 0.590 |
| Glaucoma | 0 | 19 | 16 | 60 | 0.250 | 0.168 | 0.201 |
| Glaucoma_Catarac | 0 | 12 | 18 | 65 | 0.264 | 0.684 | 0.381 |

### G.6 Gemma 10-shot ICL

| True \ Pred | Normal | Cataract | Glaucoma | Glaucoma_C | Precision | Recall | F1 |
|---|---|---|---|---|---|---|---|
| Normal | 217 | 3 | 0 | 0 | 0.660 | 0.986 | 0.791 |
| Cataract | 99 | 146 | 37 | 58 | 0.825 | 0.429 | 0.565 |
| Glaucoma | 10 | 18 | 39 | 28 | 0.375 | 0.411 | 0.392 |
| Glaucoma_Catarac | 3 | 10 | 28 | 54 | 0.386 | 0.568 | 0.460 |

### G.7 CLIP linear-probe, 10 shot

| True \ Pred | Normal | Cataract | Glaucoma | Glaucoma_C | Precision | Recall | F1 |
|---|---|---|---|---|---|---|---|
| Normal | 177 | 22 | 19 | 2 | 0.738 | 0.805 | 0.770 |
| Cataract | 50 | 56 | 70 | 164 | 0.651 | 0.165 | 0.263 |
| Glaucoma | 10 | 5 | 31 | 49 | 0.228 | 0.326 | 0.268 |
| Glaucoma_Catarac | 3 | 3 | 16 | 73 | 0.253 | 0.768 | 0.381 |

### G.8 SigLIP2 linear-probe, 10 shot

| True \ Pred | Normal | Cataract | Glaucoma | Glaucoma_C | Precision | Recall | F1 |
|---|---|---|---|---|---|---|---|
| Normal | 198 | 17 | 5 | 0 | 0.678 | 0.900 | 0.773 |
| Cataract | 77 | 66 | 36 | 161 | 0.635 | 0.194 | 0.297 |
| Glaucoma | 14 | 15 | 18 | 48 | 0.243 | 0.189 | 0.213 |
| Glaucoma_Catarac | 3 | 6 | 15 | 71 | 0.254 | 0.747 | 0.379 |

**G.9 EyeCLIP linear-probe, 10 shot**

| True \ Pred | Normal | Cataract | Glaucoma | Glaucoma_C | Precision | Recall | F1 |
|---|---|---|---|---|---|---|---|
| Normal | 44 | 88 | 0 | 88 | 0.293 | 0.200 | 0.238 |
| Cataract | 68 | 136 | 0 | 136 | 0.453 | 0.400 | 0.425 |
| Glaucoma | 19 | 38 | 0 | 38 | 0.000 | 0.000 | 0.000 |
| Glaucoma_Catarac | 19 | 38 | 0 | 38 | 0.127 | 0.400 | 0.192 |

**G.10 DINOv3 linear-probe, 10 shot**

| True \ Pred | Normal | Cataract | Glaucoma | Glaucoma_C | Precision | Recall | F1 |
|---|---|---|---|---|---|---|---|
| Normal | 190 | 13 | 5 | 12 | 0.655 | 0.864 | 0.745 |
| Cataract | 82 | 85 | 25 | 148 | 0.726 | 0.250 | 0.372 |
| Glaucoma | 12 | 12 | 18 | 53 | 0.305 | 0.189 | 0.234 |
| Glaucoma_Catarac | 6 | 7 | 11 | 71 | 0.250 | 0.747 | 0.375 |

**G.11 VisionFM (OCT) linear-probe, 10 shot**

| True \ Pred | Normal | Cataract | Glaucoma | Glaucoma_C | Precision | Recall | F1 |
|---|---|---|---|---|---|---|---|
| Normal | 97 | 87 | 21 | 15 | 0.447 | 0.441 | 0.444 |
| Cataract | 74 | 145 | 63 | 58 | 0.485 | 0.426 | 0.454 |
| Glaucoma | 26 | 34 | 14 | 21 | 0.121 | 0.147 | 0.133 |
| Glaucoma_Catarac | 20 | 33 | 18 | 24 | 0.203 | 0.253 | 0.225 |

**G.12 RETFound (OCT) linear-probe, 10 shot**

| True \ Pred | Normal | Cataract | Glaucoma | Glaucoma_C | Precision | Recall | F1 |
|---|---|---|---|---|---|---|---|
| Normal | 106 | 27 | 35 | 52 | 0.403 | 0.482 | 0.439 |
| Cataract | 105 | 25 | 84 | 126 | 0.385 | 0.074 | 0.123 |
| Glaucoma | 40 | 10 | 18 | 27 | 0.107 | 0.189 | 0.136 |
| Glaucoma_Catarac | 12 | 3 | 32 | 48 | 0.190 | 0.505 | 0.276 |

**G.13 CLIP linear-probe, full dataset**

| True \ Pred | Normal | Cataract | Glaucoma | Glaucoma_C | Precision | Recall | F1 |
|---|---|---|---|---|---|---|---|
| Normal | 41 | 3 | 0 | 0 | 0.953 | 0.932 | 0.943 |
| Cataract | 2 | 44 | 1 | 21 | 0.863 | 0.647 | 0.739 |
| Glaucoma | 0 | 3 | 7 | 9 | 0.778 | 0.368 | 0.500 |
| Glaucoma_Catarac | 0 | 1 | 1 | 17 | 0.362 | 0.895 | 0.515 |

**G.14 SigLIP2 linear-probe, full dataset**

| True \ Pred | Normal | Cataract | Glaucoma | Glaucoma_C | Precision | Recall | F1 |
|---|---|---|---|---|---|---|---|
| Normal | 41 | 3 | 0 | 0 | 0.732 | 0.932 | 0.820 |
| Cataract | 13 | 23 | 1 | 31 | 0.885 | 0.338 | 0.489 |
| Glaucoma | 2 | 0 | 1 | 16 | 0.500 | 0.053 | 0.095 |
| Glaucoma_Catarac | 0 | 0 | 0 | 19 | 0.288 | 1.000 | 0.447 |

**G.15 EyeCLIP linear-probe, full dataset**

| True \ Pred | Normal | Cataract | Glaucoma | Glaucoma_C | Precision | Recall | F1 |
|---|---|---|---|---|---|---|---|
| Normal | 44 | 0 | 0 | 0 | 0.293 | 1.000 | 0.454 |
| Cataract | 68 | 0 | 0 | 0 | 0.000 | 0.000 | 0.000 |
| Glaucoma | 19 | 0 | 0 | 0 | 0.000 | 0.000 | 0.000 |

| | | | | | | | |
|---|---|---|---|---|---|---|---|
| Glaucoma_Catarac | 19 | 0 | 0 | 0 | 0.000 | 0.000 | 0.000 |

**G.16 DINOv3 linear-probe, full dataset**

| True \ Pred | Normal | Cataract | Glaucoma | Glaucoma_C | Precision | Recall | F1 |
|---|---|---|---|---|---|---|---|
| Normal | 42 | 2 | 0 | 0 | 0.875 | 0.955 | 0.913 |
| Cataract | 6 | 36 | 4 | 22 | 0.837 | 0.529 | 0.649 |
| Glaucoma | 0 | 1 | 6 | 12 | 0.600 | 0.316 | 0.414 |
| Glaucoma_Catarac | 0 | 4 | 0 | 15 | 0.306 | 0.789 | 0.441 |

**G.17 VisionFM (OCT) linear-probe, full dataset**

| True \ Pred | Normal | Cataract | Glaucoma | Glaucoma_C | Precision | Recall | F1 |
|---|---|---|---|---|---|---|---|
| Normal | 42 | 2 | 0 | 0 | 0.737 | 0.955 | 0.832 |
| Cataract | 10 | 41 | 0 | 17 | 0.891 | 0.603 | 0.719 |
| Glaucoma | 4 | 0 | 4 | 11 | 1.000 | 0.211 | 0.348 |
| Glaucoma_Catarac | 1 | 3 | 0 | 15 | 0.349 | 0.789 | 0.484 |

**G.18 RETFound (OCT) linear-probe, full dataset**

| True \ Pred | Normal | Cataract | Glaucoma | Glaucoma_C | Precision | Recall | F1 |
|---|---|---|---|---|---|---|---|
| Normal | 27 | 16 | 0 | 1 | 0.643 | 0.614 | 0.628 |
| Cataract | 8 | 19 | 0 | 41 | 0.463 | 0.279 | 0.349 |
| Glaucoma | 6 | 4 | 0 | 9 | 0.000 | 0.000 | 0.000 |
| Glaucoma_Catarac | 1 | 2 | 0 | 16 | 0.239 | 0.842 | 0.372 |

**G.19 Bootstrapped metrics, AS-OCT classification (all sheet metrics)**

| Arm | pacg macro f1 | cataract macro f1 | cls4 macro f1 |
|---|---|---|---|
| GLM zero-shot | 0.571 (0.489-0.648) | 0.510 (0.427-0.588) | 0.335 (0.267-0.399) |
| Qwen zero-shot | 0.583 (0.495-0.671) | 0.309 (0.265-0.355) | 0.158 (0.120-0.200) |
| Gemma zero-shot | 0.615 (0.535-0.691) | 0.321 (0.272-0.375) | 0.264 (0.203-0.330) |
| GLM 5-shot ICL | 0.762 (0.694-0.822) | 0.554 (0.486-0.618) | 0.427 (0.363-0.485) |
| Qwen 5-shot ICL | 0.719 (0.651-0.782) | 0.701 (0.647-0.751) | 0.497 (0.442-0.546) |
| Gemma 5-shot ICL | 0.762 (0.701-0.814) | 0.677 (0.616-0.734) | 0.489 (0.425-0.547) |
| CLIP linear-probe, 5 shot | 0.627 (0.569-0.681) | 0.686 (0.637-0.731) | 0.443 (0.392-0.487) |
| SigLIP2 linear-probe, 5 shot | 0.658 (0.598-0.712) | 0.723 (0.670-0.772) | 0.426 (0.375-0.472) |
| EyeCLIP linear-probe, 5 shot | 0.488 (0.473-0.497) | 0.472 (0.451-0.488) | 0.214 (0.200-0.225) |
| DINOv3 linear-probe, 5 shot | 0.682 (0.617-0.740) | 0.673 (0.624-0.720) | 0.439 (0.392-0.483) |
| VisionFM (OCT) linear-probe, 5 shot | 0.583 (0.546-0.616) | 0.557 (0.536-0.576) | 0.325 (0.299-0.346) |
| RETFound (OCT) linear-probe, 5 shot | 0.500 (0.420-0.579) | 0.548 (0.513-0.580) | 0.252 (0.208-0.293) |
| VisionFM (UBM) linear-probe, 5 shot | 0.441 (0.412-0.464) | 0.450 (0.418-0.478) | 0.177 (0.156-0.194) |
| GLM 10-shot ICL | 0.750 (0.678-0.814) | 0.581 (0.513-0.644) | 0.452 (0.387-0.512) |
| Qwen 10-shot ICL | 0.727 (0.658-0.789) | 0.777 (0.719-0.829) | 0.524 (0.472-0.572) |
| Gemma 10-shot ICL | 0.780 (0.721-0.832) | 0.712 (0.654-0.764) | 0.552 (0.492-0.604) |
| CLIP linear-probe, 10 shot | 0.619 (0.555-0.681) | 0.709 (0.664-0.750) | 0.421 (0.375-0.462) |
| SigLIP2 linear-probe, 10 shot | 0.654 (0.587-0.716) | 0.716 (0.662-0.764) | 0.416 (0.370-0.456) |
| EyeCLIP linear-probe, 10 shot | 0.488 (0.473-0.497) | 0.472 (0.451-0.488) | 0.214 (0.200-0.224) |
| DINOv3 linear-probe, 10 shot | 0.670 (0.607-0.730) | 0.711 (0.660-0.758) | 0.431 (0.386-0.472) |
| VisionFM (OCT) linear-probe, 10 shot | 0.556 (0.525-0.585) | 0.549 (0.530-0.567) | 0.314 (0.288-0.336) |
| RETFound (OCT) linear-probe, 10 | 0.500 (0.418-0.578) | 0.535 (0.500-0.568) | 0.244 (0.201-0.282) |

| | | | |
|---|---|---|---|
| shot | | | |
| VisionFM (UBM) linear-probe, 10 shot | 0.443 (0.415-0.467) | 0.438 (0.406-0.465) | 0.177 (0.157-0.194) |
| CLIP linear-probe, full dataset | 0.799 (0.725-0.864) | 0.866 (0.805-0.918) | 0.674 (0.584-0.753) |
| CLIP partial-FT (last-2 layers), full dataset | 0.837 (0.765-0.899) | 0.879 (0.820-0.927) | 0.709 (0.624-0.783) |
| CLIP full-FT, full dataset | 0.871 (0.804-0.928) | 0.893 (0.839-0.940) | 0.757 (0.671-0.831) |
| SigLIP2 linear-probe, full dataset | 0.752 (0.675-0.822) | 0.771 (0.699-0.837) | 0.463 (0.391-0.535) |
| SigLIP2 partial-FT (last-2 layers), full dataset | 0.834 (0.764-0.894) | 0.800 (0.733-0.860) | 0.648 (0.563-0.725) |
| SigLIP2 full-FT, full dataset | 0.852 (0.786-0.910) | 0.912 (0.864-0.953) | 0.776 (0.700-0.843) |
| EyeCLIP linear-probe, full dataset | 0.427 (0.402-0.449) | 0.296 (0.254-0.333) | 0.113 (0.090-0.134) |
| EyeCLIP partial-FT (last-2 layers), full dataset | 0.673 (0.592-0.749) | 0.806 (0.738-0.868) | 0.508 (0.425-0.587) |
| EyeCLIP full-FT, full dataset | 0.796 (0.721-0.864) | 0.754 (0.678-0.825) | 0.552 (0.463-0.636) |
| DINOv3 linear-probe, full dataset | 0.764 (0.686-0.833) | 0.827 (0.760-0.886) | 0.604 (0.516-0.685) |
| DINOv3 partial-FT (last-2 layers), full dataset | 0.886 (0.825-0.937) | 0.883 (0.828-0.932) | 0.778 (0.697-0.849) |
| DINOv3 full-FT, full dataset | 0.890 (0.825-0.942) | 0.872 (0.814-0.923) | 0.729 (0.645-0.806) |
| VisionFM (OCT) linear-probe, full dataset | 0.795 (0.717-0.862) | 0.835 (0.771-0.893) | 0.596 (0.499-0.684) |
| VisionFM (OCT) partial-FT (last-2 layers), full dataset | 0.912 (0.853-0.960) | 0.885 (0.830-0.933) | 0.804 (0.720-0.873) |
| VisionFM (OCT) full-FT, full dataset | 0.900 (0.837-0.952) | 0.829 (0.762-0.887) | 0.739 (0.649-0.816) |
| RETFound (OCT) linear-probe, full dataset | 0.597 (0.513-0.676) | 0.714 (0.635-0.788) | 0.337 (0.277-0.393) |
| RETFound (OCT) partial-FT (last-2 layers), full dataset | 0.910 (0.853-0.957) | 0.869 (0.811-0.919) | 0.763 (0.679-0.837) |
| RETFound (OCT) full-FT, full dataset | 0.898 (0.837-0.948) | 0.875 (0.818-0.926) | 0.755 (0.668-0.831) |
| VisionFM (UBM) linear-probe, full dataset | 0.688 (0.603-0.765) | 0.793 (0.719-0.859) | 0.456 (0.395-0.513) |
| VisionFM (UBM) partial-FT (last-2 layers), full dataset | 0.837 (0.767-0.899) | 0.825 (0.759-0.882) | 0.687 (0.602-0.761) |
| VisionFM (UBM) full-FT, full dataset | 0.821 (0.748-0.884) | 0.870 (0.813-0.922) | 0.684 (0.599-0.760) |

**Figure 2, Panel H - AS-OCT Spur Localization (ASOCT-MSDC)**

Dataset size: train+val n = 956 images; test n = 150 images.

**H.1 Bootstrapped metrics, spur MAE (μm)**

| Arm | spur mae um |
|---|---|
| GLM zero-shot | 442.7 (407.3-481.3) |
| Qwen zero-shot | 733.4 (601.0-882.2) |
| Gemma zero-shot | 616.9 (573.4-665.0) |
| GLM 5-shot ICL | 402.9 (372.2-435.6) |
| Qwen 5-shot ICL | 274.1 (250.5-299.7) |
| Gemma 5-shot ICL | 550.8 (526.3-575.9) |
| CLIP linear-probe, 5 shot | 693.8 (634.8-757.8) |
| SigLIP2 linear-probe, 5 shot | 457.5 (393.9-531.5) |
| EyeCLIP linear-probe, 5 shot | 4580.2 (4549.6-4609.6) |
| DINOv3 linear-probe, 5 shot | 231.3 (212.1-251.4) |
| VisionFM (OCT) linear-probe, 5 shot | 223.2 (207.3-240.0) |

| | |
|---|---|
| RETFound (OCT) linear-probe, 5 shot | 218.9 (199.7-238.7) |
| VisionFM (UBM) linear-probe, 5 shot | 231.1 (200.8-269.0) |
| GLM 10-shot ICL | 367.7 (338.9-398.1) |
| Qwen 10-shot ICL | 273.8 (251.9-297.8) |
| Gemma 10-shot ICL | 536.2 (509.3-563.7) |
| CLIP linear-probe, 10 shot | 580.5 (530.4-636.6) |
| SigLIP2 linear-probe, 10 shot | 408.7 (358.0-475.6) |
| EyeCLIP linear-probe, 10 shot | 4666.3 (4638.7-4694.1) |
| DINOv3 linear-probe, 10 shot | 234.2 (213.7-255.4) |
| VisionFM (OCT) linear-probe, 10 shot | 212.7 (196.0-230.5) |
| RETFound (OCT) linear-probe, 10 shot | 203.5 (186.1-221.4) |
| VisionFM (UBM) linear-probe, 10 shot | 205.4 (184.2-230.8) |
| CLIP linear-probe, full dataset | 148.1 (134.3-162.6) |
| CLIP partial-FT (last-2 layers), full dataset | 148.3 (134.6-163.1) |
| CLIP full-FT, full dataset | 128.5 (115.7-142.8) |
| SigLIP2 linear-probe, full dataset | 150.5 (137.8-163.9) |
| SigLIP2 partial-FT (last-2 layers), full dataset | 150.7 (138.5-163.6) |
| SigLIP2 full-FT, full dataset | 124.7 (111.9-138.6) |
| EyeCLIP linear-probe, full dataset | 236.8 (215.5-260.2) |
| EyeCLIP partial-FT (last-2 layers), full dataset | 203.1 (185.1-221.4) |
| EyeCLIP full-FT, full dataset | 175.4 (161.3-189.9) |
| DINOv3 linear-probe, full dataset | 136.7 (123.4-151.9) |
| DINOv3 partial-FT (last-2 layers), full dataset | 137.1 (123.2-152.6) |
| DINOv3 full-FT, full dataset | 142.9 (129.0-159.0) |
| VisionFM (OCT) linear-probe, full dataset | 127.9 (115.2-141.7) |
| VisionFM (OCT) partial-FT (last-2 layers), full dataset | 133.0 (120.1-146.7) |
| VisionFM (OCT) full-FT, full dataset | 139.2 (125.5-153.5) |
| RETFound (OCT) linear-probe, full dataset | 129.5 (116.7-143.4) |
| RETFound (OCT) partial-FT (last-2 layers), full dataset | 134.8 (121.5-149.3) |
| RETFound (OCT) full-FT, full dataset | 127.8 (115.2-142.0) |
| VisionFM (UBM) linear-probe, full dataset | 131.4 (118.2-145.4) |
| VisionFM (UBM) partial-FT (last-2 layers), full dataset | 130.1 (117.3-143.6) |
| VisionFM (UBM) full-FT, full dataset | 129.2 (116.0-143.9) |

GLM spur arms use server-default reasoning: the low-effort variant substantially degraded fine-grained spur localization, so the default is retained for this panel; the variant run is archived.

**Figure 4, Panel A - Best in Classification (labelled-data scaling)**

Dataset size: per task as in Figure 2 Panels A/B/E/G; x-axis = human-annotated training images (log scale).

Each curve plots the best member of each model class (MLLM / VLM / VM) per task. Full per-arm tables: Figure 2 Panels A, B, E, G.

**F4-A.1 Best-per-class models plotted in Figure 4 Panel A**

| Task | MLLM | VLM | VM |
|---|---|---|---|
| CFP DR (QWK) | GLM5.3-Flash | CLIP | DINOv3 |
| CFP DME (QWK) | GLM5.3-Flash | CLIP | DINOv3 |
| OCT Class (macro-F1) | Qwen3.8 27B | SigLIP2 | DINOv3 |
| AS-OCT Class (macro-F1) | Gemma4-it 31B | CLIP | DINOv3 |

**Figure 4, Panel B - Best in Localization (labelled-data scaling)**

Dataset size: per task as in Figure 2 Panels C/F/H; x-axis = human-annotated training images (log scale).

Each curve plots the best member of each model class per task, selected by the full-data linear-probe endpoint. Full per-arm tables: Figure 2 Panels C, F, H.

**F4-B.1 Best-per-class models plotted in Figure 4 Panel B**

| Task | MLLM | VLM | VM |
|---|---|---|---|
| CFP vCDR (MAE, lower better) | Qwen3.8 27B | SigLIP2 | DINOv3 |
| OCT Lesion (F1) | Qwen3.8 27B | SigLIP2 | VisionFM |
| AS-OCT Spur (MAE, lower better) | Qwen3.8 27B | CLIP | VisionFM |

**Figure 4, Panel C - Cross-dataset generalization (OHTS):** the same arms as Figure 2 Panel D, plotted as REFUGE minus OHTS vCDR MAE; paired-difference CIs in Panel J.

**Figure 4, Panel C - REFUGE to OHTS cross-dataset delta (vCDR MAE)**

The delta is REFUGE MAE - OHTS MAE for the same model and arm. Every CI is a paired-difference bootstrap: each of the 10,000 replicates (seed 2026) resamples the REFUGE test images and the OHTS images independently, recomputes both pooled (micro) MAEs with the image as the resampling unit, the seed predictions collapsed per image, and invalid outputs excluded, and takes the difference. The percentile 2.5/97.5 of the replicate deltas is the CI. The loaders, validity rules and manifest ground truth are identical to the per-dataset bootstraps, so the REFUGE and OHTS columns reproduce the per-dataset supplementary tables exactly.

| Arm | REFUGE MAE | OHTS MAE | Delta (REFUGE - OHTS) | 95% CI low | 95% CI high |
|---|---|---|---|---|---|
| GLM5.3-Flash - 10-shot ICL | 0.170 | 0.150 | +0.019 | +0.009 | +0.030 |
| Gemma4-it 31B - zero-shot | 0.150 | 0.160 | -0.011 | -0.022 | +0.001 |
| Gemma4-it 31B - 10-shot ICL | 0.122 | 0.153 | -0.032 | -0.041 | -0.023 |
| GLM5.3-Flash - zero-shot | 0.104 | 0.141 | -0.037 | -0.047 | -0.028 |
| Qwen3.8 27B - zero-shot | 0.106 | 0.152 | -0.047 | -0.057 | -0.036 |
| Qwen3.8 27B - 10-shot ICL | 0.087 | 0.164 | -0.077 | -0.085 | -0.068 |
| EyeCLIP - linear probe, full pool | 0.303 | 0.369 | -0.066 | -0.121 | -0.015 |
| EyeCLIP - full fine-tune | 0.144 | 0.185 | -0.041 | -0.056 | -0.028 |
| VisionFM - linear probe, full pool | 0.115 | 0.184 | -0.069 | -0.081 | -0.058 |
| VisionFM - full fine-tune | 0.082 | 0.152 | -0.070 | -0.078 | -0.061 |
| CLIP - linear probe, full pool | 0.083 | 0.224 | -0.141 | -0.158 | -0.126 |
| CLIP - full fine-tune | 0.050 | 0.178 | -0.128 | -0.137 | -0.119 |
| SigLIP2 - full fine-tune | 0.065 | 0.278 | -0.212 | -0.225 | -0.200 |
| DINOv3 - linear probe, full pool | 0.074 | 0.223 | -0.149 | -0.161 | -0.137 |
| DINOv3 - full fine-tune | 0.075 | 0.224 | -0.149 | -0.161 | -0.138 |
| SigLIP2 - linear probe, full pool | 0.082 | 0.293 | -0.211 | -0.276 | -0.169 |
| RETFound - linear probe, full pool | 0.092 | 0.305 | -0.213 | -0.224 | -0.201 |
| RETFound - full fine-tune | 0.077 | 0.359 | -0.282 | -0.296 | -0.268 |
| EyeCLIP - fine-tune (last-2 layers) | 0.270 | 0.295 | -0.025 | -0.052 | +0.002 |
| VisionFM - fine-tune (last-2 layers) | 0.114 | 0.197 | -0.083 | -0.095 | -0.072 |
| CLIP - fine-tune (last-2 layers) | 0.064 | 0.194 | -0.130 | -0.141 | -0.120 |
| SigLIP2 - fine-tune (last-2 layers) | 0.070 | 0.216 | -0.146 | -0.157 | -0.135 |
| DINOv3 - fine-tune (last-2 layers) | 0.071 | 0.248 | -0.176 | -0.190 | -0.163 |
| RETFound - fine-tune (last-2 layers) | 0.081 | 0.380 | -0.299 | -0.312 | -0.287 |

**Figure 4, Panel D - OCT Cube Classification (WashU, Private)**

Dataset size: train+val n = 609 images; test n = 1,422 images.

**F4-D.1 GLM zero-shot**

| True \ Pred | AMD-only | DME-only | ERM-only | healthy | Precision | Recall | F1 |
|---|---|---|---|---|---|---|---|

| | | | | | | | |
|---|---|---|---|---|---|---|---|
| AMD-only | 415 | 21 | 16 | 75 | 0.933 | 0.787 | 0.854 |
| DME-only | 14 | 262 | 75 | 88 | 0.873 | 0.597 | 0.709 |
| ERM-only | 6 | 12 | 191 | 38 | 0.624 | 0.773 | 0.691 |
| healthy | 10 | 5 | 24 | 170 | 0.458 | 0.813 | 0.586 |

**F4-D.2 Qwen zero-shot**

| True \ Pred | AMD-only | DME-only | ERM-only | healthy | Precision | Recall | F1 |
|---|---|---|---|---|---|---|---|
| AMD-only | 438 | 16 | 1 | 72 | 0.907 | 0.831 | 0.867 |
| DME-only | 21 | 228 | 3 | 187 | 0.724 | 0.519 | 0.605 |
| ERM-only | 11 | 69 | 5 | 162 | 0.556 | 0.020 | 0.039 |
| healthy | 13 | 2 | 0 | 194 | 0.315 | 0.928 | 0.471 |

**F4-D.3 Gemma zero-shot**

| True \ Pred | AMD-only | DME-only | ERM-only | healthy | Precision | Recall | F1 |
|---|---|---|---|---|---|---|---|
| AMD-only | 437 | 22 | 10 | 58 | 0.783 | 0.829 | 0.806 |
| DME-only | 54 | 203 | 44 | 138 | 0.825 | 0.462 | 0.593 |
| ERM-only | 33 | 19 | 133 | 62 | 0.682 | 0.538 | 0.602 |
| healthy | 34 | 2 | 8 | 165 | 0.390 | 0.789 | 0.522 |

**F4-D.4 OCTCube-M linear-probe, 10 shot**

| True \ Pred | AMD-only | DME-only | ERM-only | healthy | Precision | Recall | F1 |
|---|---|---|---|---|---|---|---|
| AMD-only | 1073 | 1332 | 0 | 230 | 0.355 | 0.407 | 0.379 |
| DME-only | 977 | 1113 | 0 | 105 | 0.306 | 0.507 | 0.381 |
| ERM-only | 548 | 640 | 0 | 47 | 0.000 | 0.000 | 0.000 |
| healthy | 425 | 558 | 0 | 62 | 0.140 | 0.059 | 0.083 |

**F4-D.5 OCTCube-M partial-FT (last-2 layers), 10 shot**

| True \ Pred | AMD-only | DME-only | ERM-only | healthy | Precision | Recall | F1 |
|---|---|---|---|---|---|---|---|
| AMD-only | 1837 | 154 | 325 | 319 | 0.722 | 0.697 | 0.709 |
| DME-only | 262 | 1359 | 330 | 244 | 0.773 | 0.619 | 0.687 |
| ERM-only | 192 | 132 | 638 | 273 | 0.393 | 0.517 | 0.446 |
| healthy | 253 | 114 | 332 | 346 | 0.293 | 0.331 | 0.311 |

**F4-D.6 OCTCube-M linear-probe, full dataset**

| True \ Pred | AMD-only | DME-only | ERM-only | healthy | Precision | Recall | F1 |
|---|---|---|---|---|---|---|---|
| AMD-only | 328 | 199 | 0 | 0 | 0.434 | 0.622 | 0.512 |
| DME-only | 213 | 226 | 0 | 0 | 0.339 | 0.515 | 0.409 |
| ERM-only | 122 | 125 | 0 | 0 | 0.000 | 0.000 | 0.000 |
| healthy | 92 | 117 | 0 | 0 | 0.000 | 0.000 | 0.000 |

**F4-D.7 OCTCube-M partial-FT (last-2 layers), full dataset**

| True \ Pred | AMD-only | DME-only | ERM-only | healthy | Precision | Recall | F1 |
|---|---|---|---|---|---|---|---|
| AMD-only | 480 | 5 | 8 | 34 | 0.897 | 0.911 | 0.904 |
| DME-only | 11 | 407 | 5 | 16 | 0.900 | 0.927 | 0.914 |
| ERM-only | 18 | 14 | 176 | 39 | 0.903 | 0.713 | 0.796 |
| healthy | 26 | 26 | 6 | 151 | 0.629 | 0.722 | 0.673 |

**F4-D.8 Bootstrapped metrics, OCTCube-M cube**

| Arm | macro f1 | accuracy | balanced accuracy |
|---|---|---|---|
| GLM zero-shot | 0.710 (0.686-0.733) | 0.730 (0.707-0.752) | 0.743 (0.719-0.766) |
| Qwen zero-shot | 0.496 (0.476-0.516) | 0.608 (0.584-0.634) | 0.575 (0.558-0.592) |
| Gemma zero-shot | 0.631 (0.604-0.656) | 0.660 (0.635-0.684) | 0.655 (0.629-0.679) |
| OCTCube-M linear-probe, 10 shot | 0.151 (0.145-0.156) | 0.316 (0.304-0.328) | 0.243 (0.238-0.249) |
| OCTCube-M partial-FT (last-2 layers), 10 shot | 0.502 (0.489-0.513) | 0.588 (0.573-0.602) | 0.541 (0.529-0.553) |
| OCTCube-M linear-probe, full dataset | 0.230 (0.216-0.244) | 0.390 (0.364-0.416) | 0.284 (0.269-0.300) |
| OCTCube-M partial-FT (last-2 layers), full dataset | 0.822 (0.799-0.843) | 0.854 (0.835-0.872) | 0.818 (0.796-0.841) |
