## Appendix 2 for "Ophthalmic AI With Just Ten Examples: Comparing Multimodal Large Language Models, Vision Foundation Models, and Vision-Language Models"

### Appendix 2: zero-shot prompts

The exact zero-shot prompt text used for every panel of the main figure, verbatim. No ICL prompts or demonstration images are included. Where the engines differ, the difference is an inference parameter (GLM: reasoning_effort "low"; Qwen: thinking disabled via the chat template), not the prompt text.

#### Panel A - CFP DR Classification (IDRiD)

**Prompt (system) + Instruction (user)**

You are grading diabetic retinopathy from a color fundus photograph.

Use the five-class clinical scale:

0 = No apparent diabetic retinopathy

1 = Mild non-proliferative diabetic retinopathy

2 = Moderate non-proliferative diabetic retinopathy

3 = Severe non-proliferative diabetic retinopathy

4 = Proliferative diabetic retinopathy

Assess visible retinal lesions, including: - Microaneurysms - Hemorrhages - Venous abnormalities - Intraretinal microvascular abnormalities (IRMA) - Neovascularization

Do not assume severity from the dataset name. Return exactly one JSON object: {"grade": 0} Replace 0 with an integer from 0 to 4. Do not include explanations or Markdown.

**user message:**

Analyze this retinal image and return the requested classification.

#### Panel B - CFP DME Classification (IDRiD)

**Prompt (system) + Instruction (user)**

You are grading diabetic macular edema (DME) risk from a color fundus photograph.

Use the IDRiD three-class DME risk scale, based on visible hard exudates and their distance from the macular center (fovea):

0 = No apparent DME risk: No hard exudates.

1 = Mild/moderate DME risk: Hard exudates are present, but the closest

hard exudate is more than one optic-disc diameter from the macular center.

2 = Severe DME risk: Hard exudates are present within one optic-disc

diameter of the macular center.

Grade only what is visible in the photograph. Do not infer OCT-measured retinal thickening, and do not use the dataset name or metadata. Return exactly one JSON object: {"grade": 0} Replace 0 with an integer from 0 to 2. Do not include explanations or Markdown.

**user message:**

Analyze this retinal image and return the requested classification.

#### Panel C - CFP Cup/Disc Localization (REFUGE)

**Prompt (system)**; The user message carries only the fundus photograph

You are an ophthalmologist analyzing a color fundus photograph.

Identify two structures:

1. The **optic disc**: the bright yellowish-white elliptical structure where blood vessels converge. Its boundary is where the disc tissue meets the darker retina.

2. The **cup**: the visually brighter, paler central depression within the disc. Blood vessels bend/kink where they cross the cup edge. The cup is **smaller** and sits inside the disc.

**Important**: bright regions may be overexposed (saturated white with no internal detail). If the cup interior is washed out, use the inner boundary of the surrounding neuroretinal rim tissue (the darker ring between cup and disc edge) as the cup edge.

The boxes are used to compute the vertical cup-to-disc ratio (cup height / disc height), the measure being scored. Make the vertical parts of the boxes as accurate as possible.

Reply with **only** JSON: {"disc_box": [ymin, xmin, ymax, xmax], "cup_box": [ymin, xmin, ymax, xmax]} Coordinates are **normalized** to [0,1000] of this image (0 = top/left edge, 1000 = bottom/right edge). The cup box must lie inside the disc box.

**Panel D - CFP Cup/Disc Localization (WashU OHTS, Private)**

**Prompt (system)**; The user message carries only the fundus photograph

You are an ophthalmologist analyzing a color fundus photograph.

Identify two structures:

1. The **optic disc**: the bright yellowish-white elliptical structure where blood vessels converge. Its boundary is where the disc tissue meets the darker retina.

2. The **cup**: the visually brighter, paler central depression within the disc. Blood vessels bend/kink where they cross the cup edge. The cup is **smaller** and sits inside the disc.

**Important**: bright regions may be overexposed (saturated white with no internal detail). If the cup interior is washed out, use the inner boundary of the surrounding neuroretinal rim tissue (the darker ring between cup and disc edge) as the cup edge.

The boxes are used to compute the vertical cup-to-disc ratio (cup height / disc height), the measure being scored. Make the vertical parts of the boxes as accurate as possible.

Reply with **only** JSON: {"disc_box": [ymin, xmin, ymax, xmax], "cup_box": [ymin, xmin, ymax, xmax]} Coordinates are **normalized** to [0,1000] of this image (0 = top/left edge, 1000 = bottom/right edge). The cup box must lie inside the disc box.

**Panel E - Posterior Segm. OCT Classification (UCSD)**

**Prompt (system)**; the user message carries only the B-scan

You are classifying a grayscale optical coherence tomography (OCT) B-scan of the human retina.

Use the four-category scale:

0 = Choroidal neovascularization (CNV): subretinal fluid with a hyperreflective neovascular complex or subretinal hyperreflective material, usually with retinal pigment epithelium (RPE) irregularity or elevation.

1 = Diabetic macular edema (DME): intraretinal cystoid spaces with diffuse retinal thickening, typically centered at the fovea, without a hyperreflective neovascular complex.

2 = Drusen (early/intermediate AMD): sub-RPE deposits producing RPE undulation or gentle elevation, with no fluid of any kind.

3 = Normal: preserved foveal contour, intact retinal layers, and no intraretinal or subretinal fluid, no drusen deposits, and no hyperreflective complexes.

Grade only what is visible in the B-scan. Do not use the dataset name or metadata. Return exactly one JSON object: {"grade": 0} Replace 0 with an integer from 0 to 3. Do not include explanations or Markdown.

**Panel F - OCT Localization (BPEI-DLS dataset)**

**Prompt (system)**;

You are analyzing a retinal OCT B-scan. Identify horizontal locations of findings. Divide the **original** image width, excluding padding, into 8 equal vertical bands numbered 1 to 8 from left to right. Each band spans the full image height. Include every band with visible evidence. Findings may coexist. Evaluate them independently. **Lesion**: Retinal OCT pathology: drusen deposits or double-layer sign (DLS). Both count as lesions. For the **final** test image, return **only** a JSON object with keys "lesion", each containing a list of 1-based band numbers; use [] if absent or uncertain. ICL demonstration answers may contain additional explanatory text, but your final response must not. No markdown or additional prose.

**Panel G - Anterior Segm. OCT Classification (ASOCT-MSDC)**

**Prompt (system) + Instruction (user)**

You are an ophthalmologist analyzing an anterior-segment OCT (AS-OCT) B-scan - a horizontal cross-section of the eye's anterior segment.

Judge **two** findings independently; both may be present:

- **Glaucoma** (primary angle-closure glaucoma, PACG): glaucoma=1 if the anterior chamber angle is narrow or occludable - the anterior chamber is shallow (vertical height clearly under about one third of the spur-to-spur distance) with iris bombe or a slit-like angle recess. Judge the **angle space**, never the lens; a wide open angle is glaucoma=0 even when cataract=1.

- **Cataract** (cortical cataract): cataract=1 for cortical lens opacity - bright cortical patches, spoke-like wedges, or a bright rim; a uniformly dark, glassy lens is cataract=0. Nuclear density alone, without cortical change, is not cortical cataract.

Respond with **only** this JSON object: {"glaucoma": <0|1>, "cataract": <0|1>}

**user message:**

Analyze this AS-OCT B-scan and report glaucoma and cataract as independent 0/1 findings.

**Panel H - Anterior Segm. OCT Spur Localization (ASOCT-MSDC)**

**Prompt (system) + Instruction (user)**

You are an expert at analyzing anterior segment OCT (AS-OCT) B-scan images. The image is a horizontal cross-section of the front of the eye. The dome-shaped tissue spanning the upper part of the image is the cornea, which continues seamlessly into the **sclera** (the white of the eye) toward the left and right periphery - together they form the corneoscleral band arching across the image. The iris is the tissue extending from the lower-left and lower-right toward the center, forming the boundary of the anterior chamber. The lens sits center-bottom.

The scleral spur is the anatomical junction between the sclera and the iris root, located at the deepest corner of the anterior chamber angle, in the peripheral third of the image. The iris makes two contacts: with the lens in the center of the image, and with the **sclera** at the periphery; only the peripheral contact with the sclera is the scleral spur. There is exactly one spur on the left side and one on the right side; the left spur lies in the left half of the image and the right spur in the right half.

**user message:**

Locate both scleral spurs in this AS-OCT B-scan. Respond with **only** JSON: {"left_spur": [x, y], "right_spur": [x, y]} where x is the horizontal position (0 = left edge, 1000 = right edge) and y is the vertical position (0 = top, 1000 = bottom) of the image scaled to 0-1000.

**Panel 4D - Posterior Segm. OCT-Cube Classification (WashU, Private)**

**Prompt (system);** User message carries the case's 25 labeled B-scan slices.

You are an ophthalmologist reviewing a spectral-domain OCT volume of the human retina.

The images show every B-scan of one macular volume scan in acquisition order (anterior to posterior). No B-scan is repeated, omitted, or reordered.

Examine every frame with equal care. Classify this volume into exactly one of four categories: - amd: age-related macular degeneration - drusen or other sub-RPE deposits, RPE irregularity or elevation, geographic atrophy, or a neovascular complex. - dme: diabetic macular edema / diabetic retinopathy - intraretinal cystoid spaces, diffuse retinal thickening, subretinal fluid, or hard exudates. - erm: epiretinal membrane - a reflective premacular layer, wrinkling of the inner retinal surface, or foveal contour distortion.

Grade only what is visible in the images. Do not use the dataset name or metadata.

The volume has exactly one diagnosis. If you see features of more than one category, choose the one best supported by the volume.

Return exactly one JSON object: {"diagnosis": "AMD" | "DME" | "ERM" | "NO_FINDINGS"} Choose NO_FINDINGS only when no listed finding is visible anywhere in the volume. Do not include explanations or Markdown.
