## Appendix 3 for "Ophthalmic AI With Just Ten Examples: Comparing Multimodal Large Language Models, Vision Foundation Models, and Vision-Language Models"

The following sections provide additional details on the datasets, model architectures, training procedures, inference configurations, and task-specific evaluation protocols used in this study. These methods supplement the abbreviated description in the main manuscript and document the experimental procedures used to compare vision foundation models, contrastive vision–language models, and generative multimodal large language models.

### Datasets and Evaluation Tasks

Supplementary Table S1 summarizes the datasets, imaging modalities, anatomical regions, evaluation tasks, and data availability.

### Model Architectures and Inference Configurations

Supplementary Table S2 summarizes the pretrained model architectures, checkpoints, input resolutions, and adaptation approaches. For generative multimodal large language models, the table additionally reports inference configurations, including decoding parameters, reasoning settings, and maximum completion-token budgets.

### Supervised adaptation details

Vision foundation models and contrastive vision–language models were adapted using task-specific supervised prediction heads. In linear probing, pretrained image encoders were frozen and only the prediction heads were optimized. Where implemented, partial fine-tuning additionally updated selected encoder parameters, while full fine-tuning updated the complete encoder.

Few-shot experiments used 5 or 10 labeled examples per class, selected using predefined sampling manifests. For each task, shot count, and random seed, the same labeled examples were used across model families. Evaluation images were excluded from all training and demonstration sets.

Training configurations, optimization parameters, and model-selection procedures are summarized in Supplementary Table S3. Task-specific exceptions, including the volumetric OCTCube-M experiments, are described separately.

### MLLM message structure and prompting

All generative multimodal large language model (MLLM) experiments followed a standardized message-construction protocol using OpenAI-compatible message formatting. Task-specific instructions were provided in the system role and defined the clinical objective, permitted diagnostic categories or anatomical targets, and required structured JSON response format. The user role contained the test image and corresponding task request, while model-generated responses used the assistant role.

For zero-shot evaluation, models received the predefined system instruction and test image without labeled demonstrations. For in-context learning (ICL), the same system instruction was retained, and labeled image–response demonstrations were added to the conversation without updating model parameters. Demonstrations were selected using predefined sampling manifests, with 5 or 10 examples per class and no overlap with evaluation images. The message-construction framework, response-parsing procedures, and evaluation metrics were consistent across zero-shot and ICL conditions.

Model inference used a temperature of 0. Model-specific reasoning configurations and maximum completion-token budgets are reported in Supplementary Table S2. HTML renderings of the evaluation prompts were archived for reproducibility.

Task-specific implementation details

*Optic Disc and Cup Localization*

For REFUGE, vision foundation models were adapted using a convolutional segmentation head applied to frozen image-encoder representations. The head predicted pixelwise probability maps for the optic disc and optic cup and was optimized using a combination of binary cross-entropy and Dice losses. Predicted probability maps were thresholded at 0.5 to generate binary masks, from which the smallest axis-aligned bounding boxes enclosing the predicted regions were extracted. The vertical cup-to-disc ratio (vCDR) was calculated as the ratio of the predicted cup and disc bounding-box heights.

For MLLMs, optic disc and cup localization was performed through direct bounding-box prediction using standardized task instructions. Predicted bounding-box dimensions were used to calculate vCDR. Cross-dataset generalization was assessed on OHTS using REFUGE-derived labeled demonstrations, without using OHTS images as in-context examples.

*Anterior-Segment OCT Evaluation*

Anterior-segment OCT (AS-OCT) images were evaluated for two tasks: binary classification of anatomical angle closure versus non-angle closure and localization of the scleral spur. Classification performance was assessed using standard classification metrics.

Localization performance was assessed using Euclidean distance between predicted and reference landmark coordinates, converted to micrometers (1 pixel = 7.4941 μm, Tomey CASIA2 16-mm scan width), reported as the image-level median with 95% bootstrap CIs. For zero-shot MLLMs, the scleral spur was defined in the instructions as the termination point of the corneoscleral band at the deepest corner of the angle recess, and coordinates were returned on a 0–1000 normalized scale. For in-context learning, demonstrations showed the scleral spur painted as a magenta blob and the model returned left and right spur coordinates. Supervised vision models localized the spur with segmentation heads trained on Gaussian-smoothed landmark maps centered on the annotated spur points, with the predicted location taken from the peak of the predicted heatmap.

*Band-Level Retinal Lesion Localization*

For the internal retinal OCT dataset, each B-scan was divided into eight vertical bands to evaluate the spatial localization of annotated retinal lesions. A band was considered positive when it contained at least four annotated lesion pixels.

For MLLMs, Set-of-Marks (SoM) prompting was used to provide explicit spatial references. Numeric markers were overlaid on the eight bands, allowing models to identify lesion-containing regions by their corresponding band numbers. The same band definitions were used across model families, and localization performance was assessed using band-level F1 score and balanced accuracy.

### Occlusion Sensitivity Analysis

Overview and image selection. Occlusion sensitivity analyses were conducted on retinal optical coherence tomography (OCT) images from UCSD and color fundus photographs (CFP) from IDRiD to examine the spatial dependence of model predictions. Representative correctly classified images were selected for qualitative comparison, and visualization examples are shown in Figure 3a (CFP DR classification) and Figure 3d (OCT classification). For UCSD, the analysis included images from four diagnostic categories: choroidal neovascularization (CNV), diabetic macular edema (DME), drusen, and normal retina. For IDRiD, the analysis focused on five-class diabetic retinopathy (DR) severity classification.

Zero-shot classification and class-evidence scoring. Occlusion analyses were performed using the same zero-shot classification instructions and model-specific text prompts as the corresponding classification evaluations.

For Qwen3.8-27B, the original system and user instructions were used with reasoning disabled. In the UCSD classification task, the model was instructed to assign one of four numeric labels: 0 (CNV), 1 (DME), 2 (drusen), or 3 (normal), and to return the prediction as a JSON object containing the key grade. During the original classification evaluation, Qwen generated its JSON response autoregressively. For occlusion sensitivity analysis, class-specific evidence was instead extracted directly from next-token logits. The model received the same system instruction, user instruction, and image as in the original evaluation, with an additional assistant response prefix, {"grade": . The logits corresponding to the four candidate numeric labels (0, 1, 2, and 3) were extracted without generating a complete response. Each numeric label was verified to correspond to a single tokenizer token (token IDs 15, 16, 17, and 18, respectively). Softmax normalization over these four candidate logits produced a closed-set distribution of relative class preferences. This prefix-based scoring differs from the original autoregressive classification procedure because it constrains the next-token position to the numeric grade field. However, the prefix specifies only the response structure already required by the original classification prompt and introduces no additional diagnostic information. The resulting scores were therefore interpreted as conditional class preferences rather than probabilities of freely generated responses.

For contrastive vision–language models, class-specific evidence was derived from image–text similarity. Image embeddings were compared with text embeddings representing the diagnostic categories, using the same prompt configuration and model-specific similarity scaling as in the zero-shot classification evaluation. The resulting class logits were normalized using softmax across the candidate classes. For EyeCLIP, cosine similarities between normalized image and text embeddings were multiplied by a fixed scaling factor of 100. Softmax normalization was subsequently applied across the candidate classes. For SigLIP2, similarity scores were transformed using the model's learned logit scale and bias. The learned SigLIP2 bias is shared across the class logits and therefore cancels under softmax normalization. Although these scaling procedures do not change the class ranking within an image, they influence the resulting probability distributions and the magnitude of occlusion sensitivity scores. Accordingly, the original model-specific scaling procedures were retained.

The class-evidence scores obtained from generative and contrastive models are not directly equivalent: Qwen produces next-token logits over its vocabulary, whereas contrastive models produce image–text similarity scores. Restricting the scores to the predefined diagnostic categories and applying softmax provides a common representation of relative class preference. Nevertheless, differences in score calibration across architectures remain, and occlusion effects were interpreted within each model rather than as directly comparable measures of absolute disease evidence.

Occlusion procedure. Each image was divided into an 8 × 8 grid comprising 64 spatial regions. Each region was sequentially replaced with the median pixel intensity of the original image while all remaining regions were preserved. For each perturbed image, class-evidence scores were recalculated using the same model, text prompts, preprocessing, and scoring procedure as for the unoccluded image.

Positive sensitivity values indicated regions whose removal reduced the model's relative preference for the target class, whereas negative values indicated regions whose removal increased that preference.

Task-specific text prompts. For UCSD, Qwen used the original OCT classification prompt specifying the four diagnostic categories and their corresponding imaging characteristics. The prompt described CNV in terms of subretinal fluid and neovascular complexes, DME in terms of intraretinal cystoid spaces and retinal thickening, drusen in terms of sub-retinal pigment epithelium deposits without fluid, and normal retina in terms of preserved retinal anatomy without pathological findings. The accompanying user instruction was: “Analyze this OCT B-scan and return the requested classification”. For EyeCLIP and SigLIP2, class-specific OCT descriptions were used as text prompts:

- CNV: “A retinal OCT B-scan showing choroidal neovascularization with subretinal fluid and a neovascular complex.”
- DME: “A retinal OCT B-scan showing diabetic macular edema with intraretinal cystoid spaces.”
- Drusen: “A retinal OCT B-scan showing drusen, sub-RPE deposits, and RPE undulation without fluid.”
- Normal: “A normal retinal OCT B-scan with intact retinal layers and no pathology.”

An alternative configuration using only diagnostic class names (“choroidal neovascularization,” “diabetic macular edema,” “drusen,” and “normal”) was also examined to assess sensitivity to text-prompt formulation.

For IDRiD, the contrastive model prompts represented the five DR severity categories: “no diabetic retinopathy,” “mild nonproliferative diabetic retinopathy,” “moderate nonproliferative diabetic retinopathy,” “severe nonproliferative diabetic retinopathy,” and “proliferative diabetic retinopathy”.

#### WashU OCT Cohort Construction

*Motivation*

Clinical OCT examinations consist of multiple B-scans, and pathological findings may be localized to a subset of the volume; a single representative B-scan may therefore not characterize the examination. To test whether a multimodal large language model (MLLM)-based inference can operate under this real-world condition, we constructed an enriched, single-finding cohort of EHR-linked Spectralis OCT examinations from Washington University in St Louis.

*Study population and data sources*

We identified macular OCT volumes acquired with Spectralis (Heidelberg Engineering) between January 2013 and October 2026. Each volume was linked to the patient's ophthalmology encounters in the electronic health record, including billed diagnoses and clinical notes. Of 199,486 imaging series, 151,165 were macular volumes with at least 25 B-scans, and 129,381 of these could be linked to an ophthalmology encounter on the day of imaging or within 30 days afterward. The 30-day window reflects the common workflow in which imaging is obtained at one visit and the diagnosis is recorded at the follow-up visit.

*Class definitions and eligibility*

The four classes were AMD, DME, ERM, and healthy. Disease classes were defined from ICD-10 diagnoses. To create single-disease classes, the diagnoses at the linked encounter had to indicate exactly one of the three diseases, and the patient's coded history could contain no other retinal disease, including diabetic retinopathy without macular edema, macular hole, retinal detachment, and retinal vascular disease. Eyes with confounding ocular diagnoses on the day of imaging (vitreous hemorrhage, endophthalmitis or panuveitis, optic nerve disease, iridocyclitis, ocular tumors, congenital posterior-segment anomalies) were excluded. Treatment histories inconsistent with the assigned class were exclusionary, such as anti-VEGF injection for ERM and any retinal treatment for healthy eyes. This single-finding design responds to the structure of real-world coding, in which multiple retinal diagnoses frequently co-occur within one encounter, and isolates each finding's imaging signature at the cost of excluding co-positive volumes.

*Eye-specific labeling*

Because patients are often imaged in one eye while carrying a diagnosis in the other, labels were assigned per eye. The laterality of each diagnosis was taken from the clinician-entered diagnosis description (eg, "right eye," "bilateral") or, when the description did not specify, from the laterality encoded in the ICD-10 code; when the two disagreed, the clinician-entered description took precedence. A diagnosis contributed to a volume's label only if it referred to the imaged eye or to both eyes; fellow-eye and unspecified diagnoses were not used. A volume was labeled with a disease only if this eye-specific evidence indicated exactly one disease and the imaged eye's record contained no other retinal disease at any time.

*Clinical note review*

Because billing codes can miss findings documented in the chart, we screened the assessment and plan of each candidate's ophthalmology notes with a rule-based keyword algorithm that classified every statement as affirmative (eg, "drusen present"), negated (eg, "no ERM"), or uncertain/family-history-related (eg, "possible drusen"; "father with AMD"). Only affirmative statements counted as evidence of disease. Negated statements were treated as evidence of disease absence and could therefore contradict a coded label, but never affirmed one; uncertain and family-history mentions carried no evidentiary weight in either direction. The vocabulary and rules were reviewed by a retina specialist. Volumes were excluded when the notes documented a second study disease or contradicted the assigned label.

*Cohort selection*

Of the linked volumes, 49,917 from patients with AMD, DME, or ERM met the single-disease criteria on initial screening of patient-level diagnosis codes. Because patients with chronic retinal disease are imaged repeatedly, these volumes came from fewer than 3,000 patients; we therefore retained one volume per patient (the most recent qualifying examination), so that repeated scans of the same eye were not counted as independent samples. After treatment-based and pathology exclusions and the addition of healthy controls, 2,576 candidate volumes remained (886 AMD, 777 DME, 578 ERM, 335 healthy).

Clinical note review and eye-specific labeling then excluded 545 volumes (134 AMD, 150 DME, 225 ERM, 36 healthy).

*Final dataset and partitions*

The final dataset comprised 2,031 OCT volumes from 2,031 patients: 752 AMD, 627 DME, 353 ERM, and 299 healthy. Volumes were partitioned at the patient level using stratified random sampling (seed 42) into a held-out test set (70%; n = 1,422: 527 AMD, 439 DME, 247 ERM, 209 healthy) and a development pool (30%; n = 609), the latter further divided into training (n = 426) and validation (n = 183) subsets for fully supervised experiments. Few-shot demonstration sets (10 patients per class; 40 volumes total) were generated for five seeds (42–46) exclusively from the training subset. Split integrity was verified programmatically: no patient overlap across training, validation, and test; no few-shot overlap with validation or test; no duplicate image identifiers; no conflicting patient labels; all cases accounted for.

### Volumetric OCT Model Evaluation

*Zero-Shot MLLM Evaluation*

Three generative MLLMs (Qwen3.8-27B, GLM-5.3-Flash, and Gemma-4-31B-it) were evaluated for zero-shot four-class classification of clinical OCT volumes into age-related macular degeneration (AMD), diabetic macular edema (DME), epiretinal membrane (ERM), or no findings.

Each OCT volume was represented by 25 B-scans encoded as JPEG images. All 25 images were provided together in a single multimodal inference request, allowing the model to generate one volume-level classification. Models received a standardized clinical instruction specifying the diagnostic categories and required JSON response format. No task-specific labeled demonstrations or parameter updates were used.

*Supervised OCTCube-M Evaluation*

As a supervised volumetric comparator, OCTCube-M, a three-dimensional OCT foundation model pretrained using masked autoencoding, was initialized from its official pretrained checkpoint. Each OCT volume was resized to 60 × 256 × 256 voxels using trilinear interpolation to match the pretrained model's expected input dimensions.

Few-shot adaptation used 10 labeled volumes per class, selected exclusively from the training subset across five random seeds (42–46). Two adaptation strategies were evaluated: linear probing, in which the pretrained encoder remained frozen and only the classification head was trained, and partial fine-tuning, in which the final two transformer blocks and classification head were updated.

Models were trained for 30 epochs, and the checkpoint achieving the best validation performance was selected using the predefined validation subset. Final classification performance was evaluated on the same held-out test set used for MLLM evaluation.

The volumetric experiment therefore compared zero-shot MLLM inference, requiring no task-specific labeled examples, with supervised OCTCube-M adaptation using a limited number of labeled training volumes and a separate validation set. Unlike the matched few-shot experiments conducted on the other benchmark datasets, the volumetric comparison did not use equivalent labeled-data budgets or adaptation procedures.
