## Supplemental Table 1 for "Ophthalmic AI With Just Ten Examples: Comparing Multimodal Large Language Models, Vision Foundation Models, and Vision-Language Models"

**Supplementary Table S1. Ophthalmic imaging datasets and evaluation tasks**

| Dataset | Anatomical region | Modality | Evaluation task | Availability |
|---|---|---|---|---|
| UCSD OCT | Posterior segment (retina) | OCT | Four-class disease classification: normal, CNV, DME, drusen | Public |
| IDRiD | Posterior segment (retina) | CFP | Diabetic retinopathy (DR) severity grading (grades 0–4); DME risk grading (grades 0–2) | Public |
| REFUGE | Posterior segment (optic nerve head) | CFP | Optic disc and cup localization and vertical cup-to-disc ratio (vCDR) estimation; source of labeled demonstrations for MLLM in-context learning on OHTS. | Public |
| OHTS | Posterior segment (optic nerve head) | CFP | External validation of vertical cup-to-disc ratio (vCDR) estimation using few-shot demonstrations selected from REFUGE. | Controlled access |

| | | | | |
|---|---|---|---|---|
| BPEI-DLS dataset | Posterior segment (retina) | OCT | Band-level lesion localization: drusen, double-layer sign (DLS), geographic atrophy (GA) | Private |
| AS-OCT dataset | Anterior segment (iridocorneal angle) | AS-OCT | Multi-label classification of PACG and cataract status (including binary PACG and cataract classification and four-class categorization); scleral-spur landmark localization. | Public |
| WashU clinical dataset | Posterior segment (retina) | OCT volumes + infrared en face fundus photograph | EHR-linked four-class classification of AMD, DME, ERM, and healthy eyes using volumetric OCT. | Private |

**Abbreviations:**

**Datasets:** UCSD, University of California, San Diego; IDRiD, Indian Diabetic Retinopathy Image Dataset; REFUGE, Retinal Fundus Glaucoma Challenge; OHTS, Ocular Hypertension Treatment Study; BPEI, Bascom Palmer Eye Institute; WashU, Washington University in St. Louis.

**Imaging modalities:** AS-OCT, anterior-segment optical coherence tomography; CFP, color fundus photography; OCT, optical coherence tomography.

**Diagnoses and findings:** AMD, age-related macular degeneration; CNV, choroidal neovascularization; DME, diabetic macular edema; DLS, double-layer sign; DR, diabetic retinopathy; ERM, epiretinal membrane; GA, geographic atrophy; PACG, primary angle-closure glaucoma; vCDR, vertical cup-to-disc ratio.

**Other:** EHR, electronic health record; MLLM, multimodal large language model.
