## Supplemental Table 2 for "Ophthalmic AI With Just Ten Examples: Comparing Multimodal Large Language Models, Vision Foundation Models, and Vision-Language Models"

**Supplementary Table S2. Model architectures, checkpoints, and inference configurations.** Panel A summarizes the evaluated model families, pretrained checkpoints, input image resolutions, and downstream adaptation approaches. For EyeCLIP, we used the official revised pretrained checkpoint (eyeclip.pt), released in February 2026. Panel B summarizes generation settings for MLLMs. All models used temperature zero, with explicitly specified completion-token limits. Thinking was disabled for Qwen3.8-27B and Gemma-4-31B-it, whereas GLM-5.3-Flash used low reasoning effort with thinking enabled.

**Panel A. Model architectures, pretrained checkpoints, and adaptation strategies.**

| Model | Category | Checkpoint / pretraining | Input resolution | Adaptation |
|---|---|---|---|---|
| DINOv3 | VM (self-supervised) | facebook/dinov3-vitb16-pretrain-lvd1689m | 448 × 448 (positional interpolation) | Linear probe (frozen backbone); last-2 fine-tuning at full data |
| RETFound | VM (domain-pretrained) | OCT: RETFound_mae_natureOCT.pth; CFP: RETFound_mae_natureCFP.pth; | 448 × 448 | Linear probe (frozen backbone); last-2 fine-tuning at full data |
| VisionFM | VM (medical-pretrained) | OCT: VFM_OCT_weights.pth CFP: VFM_Fundus_weights.pth UBM: VFM_UBM_weights.pth | 448 × 448 | Linear probe (frozen backbone); last-2 fine-tuning at full data |
| CLIP | Contrastive VLM | openai/clip-vit-large-patch14 | 448 × 448 (positional interpolation) | Zero-shot image–text similarity; linear probe |

| | | | | |
|---|---|---|---|---|
| EyeCLIP | Contrastive VLM (ophthalmic) | Official EyeCLIP ViT-B/32; local eyeclip.pt checkpoint | 448 × 448 | Zero-shot image–text similarity; linear probe |
| SigLIP2 | Contrastive VLM | google/siglip2-base-patch16-512 | 512 × 512 | Zero-shot image–text similarity; linear probe |
| Qwen3.8-27B | MLLM | Qwen/Qwen3.8-27B | Dynamic | Zero-shot / few-shot ICL (no parameter updates) |
| GLM-5.3-Flash | MLLM | nvidia/GLM-5.3-Flash-NVFP4 | Dynamic | Zero-shot / few-shot ICL (no parameter updates) |
| Gemma-4-31B-it | MLLM | google/gemma-4-31B-it | Dynamic | Zero-shot / few-shot ICL (no parameter updates) |

**Panel B. Generative MLLM inference parameters**

| Parameter | Qwen3.8-27B | GLM-5.3-Flash | Gemma-4-31B-it |
|---|---|---|---|
| Serving framework | vLLM | vLLM | vLLM |
| Temperature | 0 | 0 | 0 |

| | | | |
|---|---|---|---|
| Maximum completion tokens | 4,096 | 8,192 | 4,096 |
| Thinking mode | Disabled | Enabled | Disabled |
| Reasoning effort | Not applicable | Low (AS-OCT spur localization exception) | Not applicable |
| Reasoning parser | qwen3 | glm45 | gemma4 |
| Top-p (model configuration) | 0.95 | 0.95 | 0.95 |
| Top-k (model configuration) | 20 | Not specified | 64 |
| Output format | Prompt-enforced structured JSON | Prompt-enforced structured JSON | Prompt-enforced structured JSON |
| Maximum server context length | 32,768 | 1,048,576 | 32,768 |
| Model weight precision/quantization | BF16 | NVFP4 | BF16 |
| KV cache precision | FP8 | FP8 | FP8 |

**Abbreviations:**

**Model architectures and methods:** VM, vision model; VLM, vision–language model; MLLM, multimodal large language model; ICL, in-context learning; ViT, Vision Transformer; CLIP, Contrastive Language–Image Pretraining; DINO, self-distillation with no labels; RETFound, retinal foundation model; SigLIP, sigmoid loss for language–image pretraining; GLM, General Language Model.

**Imaging modalities:** OCT, optical coherence tomography; CFP, color fundus photography; UBM, ultrasound biomicroscopy; AS-OCT, anterior-segment optical coherence tomography.

**Inference and computational settings:** BF16, brain floating point 16-bit; FP8, 8-bit floating point; NVFP4, NVIDIA 4-bit floating-point format; KV, key–value; JSON, JavaScript Object Notation.
