## Supplemental Table 3 for "Ophthalmic AI With Just Ten Examples: Comparing Multimodal Large Language Models, Vision Foundation Models, and Vision-Language Models"

**Supplementary Table S3. Training hyperparameters and model selection procedures for supervised adaptation.**

| Parameter | Value |
|---|---|
| Optimizer | AdamW |
| Classification-head learning rate | $1 \times 10^{-3}$ |
| Backbone learning rate (fine-tuning modes) | $1 \times 10^{-5}$ |
| Weight decay | 0.01 |
| Gradient clipping | 1.0 |
| Precision | BF16 autocast |
| Few-shot epochs | 10 for classification; 30 for specified localization tasks; fixed, without validation-based selection or early stopping |
| Full-data epochs | 30, with internal-validation best-epoch selection |
| Class imbalance handling | Positive-class weighting in the loss |
